# A Regionally Determined Climate-Informed West Nile Virus Forecast Technique

**DOI:** 10.1101/2025.03.27.25324789

**Authors:** Ryan D. Harp, Karen M. Holcomb, Stanley G. Benjamin, Benjamin W. Green, Hunter Jones, Michael A. Johansson

**Affiliations:** Global Systems Laboratory, National Oceanic and Atmospheric Administration; Boulder, CO, USA; Division of Vector-Borne Diseases, Centers for Disease Control and Prevention; Fort Collins, CO, USA; Cooperative Programs for the Advancement of Earth System Science, University Corporation for Atmospheric Research; Boulder, CO, USA; Physical Sciences Laboratory, National Oceanic and Atmospheric Administration; Boulder, CO, USA; Cooperative Institute for Research in Environmental Science, University of Colorado Boulder; Boulder, CO, USA; Climate Program Office, National Oceanic and Atmospheric Administration; Silver Spring, MD, USA; Division of Vector-Borne Diseases, Centers for Disease Control and Prevention; San Juan, PR, USA; Network Science Institute and Bouvé College of Health Sciences, Northeastern University; Boston, MA, USA

## Abstract

**Background:** West Nile virus (WNV) infection has caused over 30,000 human cases of the severe, neuroinvasive form of the disease (West Nile virus Neuroinvasive Disease; WNND) and nearly 3,000 deaths in the U.S. since its introduction in 1999. Despite spatiotemporal variation in the impact of WNV and its known links to various climate factors, no effective nationwide WNV or WNND forecast exists.

**Objectives:** We aim to produce a skillful, nationwide WNND forecast built upon regionally varying relationships between climate factors and WNND.

**Methods:** We examined the impact of climate conditions on annual WNND caseload for 11 ecologically meaningful regions in the U.S. The most salient climate factors were incorporated into a regionally determined nationwide WNND forecast model. We retrospectively generated forecasts from 2005-2022 using observed climate conditions and compared forecast skill against various benchmarks, including a simple, historical case-driven model. Forecast skill was assessed by weighted interval scoring.

**Results:** Regional, climate-informed WNND retrospective forecasts outperformed a benchmark model only informed by historical WNND case data across all regions, as well as in a nationally aggregated score (univariate: 18.1% [3.7–27.0%], bivariate: 23.9% [9.0–32.8%]). Additionally, the regional forecasts outperformed an ensemble model generated from the 2022 CDC WNV Forecasting Challenge and a parallel, county-level, regional climate-informed forecast outperformed forecasts from the same Challenge. Drought and temperature were the climate factors most consistently linked to WNND and incorporated into our forecast model.

**Discussion:** We show a retrospectively generated WNND forecast for the continental U.S. that considerably improved upon simple forecasts based on historical case distributions. This forecast aggregated county-level data to broader regions to boost statistical signal and capture the regionally varying influences of climate conditions on annual WNND caseload. The advances here represent a potential path toward actionable broad-scale WNV forecasts.

## Introduction

West Nile virus (WNV) arrived in the U.S. in 1999 and has since become the most prevalent mosquito-borne disease in the continental U.S.^1,2^. While most WNV cases are asymptomatic (75–80%)^3,4^, about 1-in-150 WNV infections lead to a severe, neuroinvasive form of the disease known as West Nile neuroinvasive disease (WNND). Ten percent of WNND cases are fatal and many who survive have enduring disabilities as a result^5,6^. Through the end of 2023, over 30,000 cases of WNND and nearly 3,000 WNV-caused deaths have been reported nationally^7^.

In the U.S., WNV is vectored by *Culex* mosquitoes and amplified in avian hosts, predominantly *passerines* (e.g., songbirds). WNV is thus intrinsically coupled to the environmental conditions that affect its host and vector species and has been linked to a large number of climate variables, including temperature, drought, and precipitation^8,9^. However, the relationships between climate factors and WNV and its vectors are complex. For example, while warm temperatures can lead to quickened development of mosquitoes and increased rates of WNV amplification and transmission, this relationship is constrained by the biological thermal limit of mosquitoes and temperatures that are too warm can lead to increased mosquito fatality^10,11^. Precipitation can have similar competing effects on mosquito abundance. While increased precipitation can lead to expanded breeding habitat and an increased mosquito population in water-limited locations^12,13^, high-intensity precipitation can wash away existing breeding sites, including the juvenile mosquitoes developing in those sites, and limit mosquito populations^14,15^. These relationships are also regionally dependent^12,16,17^. While drought has been shown to increase WNV transmission^9^, the mechanistic links between drought and WNV are particularly intricate. Decreased mosquito predation, an increased frequency of contact between vectors and hosts at a reduced number of water sources, and altered mosquito behavior driving them toward artificial water sources that are closer to people, have all been cited as reasons why WNV transmission is increased by drought^9,18,19^. Complex relationships like these lead to regional differences in the influence of climate conditions on WNV transmission. Additional climate variables that have been shown to affect WNV transmission include relative humidity, dew point, vapor pressure deficit, solar radiation, day length, soil moisture, and snow conditions^9,12,20–22^.

Due to the high impact of WNV in the U.S., there is strong interest in generating skillful nationally scoped WNV forecasts to aid proactive public health responses like vector control and WNV awareness campaigns^2^. However, despite this interest and the known links between environmental conditions and WNV that could be leveraged toward WNV forecasts, no effective nationwide WNV forecast exists^23–25^.. Note that here we define a “nationwide” or “national” forecast as one that collectively covers the majority of the country, regardless of the scale of the forecast target (i.e., a nationwide forecast could be comprised of many county-, state-, or region-specific forecasts and does not imply a singular national estimate of WNV caseload). Recent county-level WNV Forecasting Challenges led by the U.S. Centers for Disease Control and Prevention (CDC) found that basic disease forecasts informed solely by distributions of historical WNND caseloads (i.e., not using the year-over-year sequence of cases) are among the most skilled nationwide WNV forecasts. Despite these simple forecasts ultimately generating baselines that do not tailor projections to the forecast-year, they consistently outperformed more sophisticated forecasting models^23,25^. These results, as well as inconclusive model covariate analyses regarding the inclusion of climate factors in these Challenges, suggest that climate information has not been appropriately incorporated into WNV disease forecast models.

Several potential reasons exist for this lack of success. One barrier is the infrequency with which WNND occurs at the county level^7^, making extracting statistical signals at the annual-county scale difficult. Of all county-years under consideration, 91% did not report a single case of WNND and < 1% of county-years reported more than one case of WNND. Further, while interannual variability in climatic conditions can drive differences in annual WNND caseload, the connections between climate and WNV are heterogeneous due to underlying differences in local ecology and climatic conditions, making the creation of a nationwide forecast complicated^26^. The complexity of the links between climate and WNV further muddle forecast development. However, despite the lack of success and inherent difficulties in producing effective nationwide WNV forecasts, some local forecasts have shown promising results in forecasting WNV or its vectors (New York^27,28^, California^29,30^, South Dakota^21^). Along with factors like increased data availability at a more focused scale, these localized forecasts may have benefited from relatively consistent ecology (i.e., vector species and their predators) given the more constrained forecast area and, consequently, more consistent relationships between environmental factors and WNV^26^. Their success provides evidence that appropriately scaled approaches–informed by intrinsic ecological and climatic spatial variability and dependent on the outcome under consideration and data availability– may prove effective.

Here, we develop a novel, regionally based approach to WNV forecasting that is informed by local climate conditions. This approach aims to overcome two critical issues in developing large-scale forecasts. First, we employ regional aggregation to combat limitations in statistical signal driven by sparse county-level WNND caseload. Second, we determine the climate drivers of our forecast model at a regional level to allow for discrepancies in the influence of climate on WNND that may be driven by differences in local vector and host species and underlying climatic conditions. The remainder of this manuscript describes our approach, characterizes its effectiveness using historical data, and details lessons learned.

## Methods

### WNND and Climate Data

We centered our analysis on WNND cases–as opposed to all WNV cases–as WNND cases are the most likely to be properly diagnosed and consistently reported due to the severity of the disease (∼1-in-150 of all WNV cases^5^), though we subsequently refer to forecasts as WNV forecasts given that WNND is representative of overall WNV burden. We used county-annual WNND data from 2005-2022 that is publicly available from ArboNET, the national arboviral disease surveillance system administered by CDC^7^. While WNND data was available beginning in 1999, we began our analysis in 2005, the year after WNV had completed its initial epidemic across the continental U.S. We combined this data with climate data from multiple historical datasets, including PRISM^31^, gridMET^32^, and NLDAS^33^. PRISM (Parameter-elevation Regressions on Independent Slopes Model) and gridMET (Gridded Surface Meteorological) are both observationally based, gridded near-surface products statistically downscaled to roughly 4-km of resolution^31,32^. NLDAS (National Land Data Assimilation System) is a data assimilation product that merges a wide array of observations and model reanalysis data via land surface and hydrological models to produce output at roughly 9–14-km (⅛ degree latitude/longitude) of resolution across the continental U.S.^33^. We included the following variables in our exploratory analysis: monthly values of 2-m mean temperature (i.e., temperature at two meters above ground level), 2-m dew point, precipitation, vapor pressure deficit maximum (PRISM), Palmer Drought Severity Index (PDSI; gridMET), and soil moisture at 50-mm depth (NLDAS).

### NEON Regions

To combat the statistical limitations posed by low WNND case counts at the county scale, we aggregate WNND and climate data onto larger ecologically meaningful and climatically informed spatial regions determined by the National Ecological Observatory Network (NEON). NEON defined 17 regions around the continental U.S. through a multivariate clustering method built from nine ecologically relevant climate variables^34^. This regional aggregation approach also ameliorates issues posed by the spatiotemporal mismatches between disease data and climate data by moving analysis away from administrative boundaries (e.g., counties or states) onto regions with internally consistent but regionally varying climate baselines and variability.

While our analysis was performed at the regional level, we first spatially averaged our climate data at the county level using county boundary shapefiles available from the U.S. Census Bureau^35^. Counties were then classified to the appropriate NEON regions based on the location of their centers of population^36^; and WNND counts were summed for each region^37^ (Figure S1). We limited our analysis here to the 11 regions that had reported a minimum of 1,000 cases of WNND; these regions had collectively reported > 93% of all U.S. WNND cases. To more accurately reflect the relationship between climate and WNND caseload in each region, we create weighted averages of climate time series–as opposed to simple spatial averages–by weighting county-level climate data by the natural log of reported historical WNND cases. Counties without reported WNND cases were not included. This method emphasizes the climatic conditions for the locations of most impact (i.e., high WNND caseload counties) and allows for a more effective characterization of the influence of climate on the interannual variability of WNND.

### Investigating the Influence of Climate on WNND

Given the widespread variability in *Culex* mosquito habitat suitability and abundance by species^38,39^, as well as the large spatial differences in the climatic factors that affect WNV vectors^17^, it is critical to allow for spatial heterogeneity in the relationships between climate and WNV. With this motivation, we performed exploratory analyses by assessing Pearson correlations between natural logged annual WNND case counts and each month-variable combination individually (e.g., March PDSI, May mean temperature) for all regions. This allowed us to identify the climate factors that most affect WNND cases for each region. Due to the strong seasonality of WNV in the U.S. (an August–September peak accounts for 74% of all WNND cases with 96% of WNND cases occurring between July–October), we compared annual WNND case counts to climate conditions from the previous October through the concurrent July.

### Producing Probabilistic Forecasts

We used negative binomial generalized linear regression models in a Bayesian framework to produce probabilistic regional-level forecasts of annual WNND case counts (*rstanarm* package in *R*^40,41^). This technique allows for modeling discrete outcomes that may be highly dispersed, such as WNND case counts, and the inclusion of multiple predictors. We produced independent forecasts for each of the 11 NEON regions with moderate to high historical WNND caseload: Northeast, Mid Atlantic, Southeast, Great Lakes, Prairie Peninsula, Ozarks Complex, Northern Plains, Central Plains, Southern Plains, Desert Southwest, and Pacific Southwest (Figure S1).

Based on the exploratory correlation analysis, we used the monthly climate variables with the strongest correlation with WNND to inform both a univariate and bivariate model (Table 1, Figures S2-12). This was done independently for each region to allow for regional variation in the importance of climate factors on WNV and its vectors. Because of the strong seasonal peak in WNV occurrence in August–September and the inherent temporal lag in the influence between climate conditions and WNV cases, we restricted our included variables to those in July or earlier. To reduce autocorrelation between our selected input climate variables, we limited selection of the secondary climate variable to those of a separate grouping of interrelated variables from that of the initial variable (near-surface air conditions: mean temperature, dew point, vapor pressure deficit max; antecedent soil moisture conditions: PDSI, 50-mm soil moisture; and precipitation). The selected regional-level climate variables (Table 1) were then regressed upon annual WNND counts assuming a negative binomial distribution of raw WNND cases for each region (i.e., cases were not log-transformed) using noninformative default priors (Text S1). To create a probabilistic forecast from these Bayesian regressions, we generated a series of 23 quantiles (1%, 2.5%, 5%, 10%, 15%, … 85%, 90%, 95%, 97.5%, 99%) from the posterior predictive distribution to mirror the format used in the 2022 CDC West Nile Virus Forecasting Challenge^2^.

**Table 1:** Climate Variables Used in Forecast Model Regression. The month-climate variable combinations used as inputs for the univariate (primary climate factors) and bivariate (primary and secondary climate factors) regional WNV forecast models, along with the Pearson correlation coefficients between the primary and secondary climate factors for each region.

| NEON Region | Primary Climate Factors | Secondary Climate Factors | Correlation between Climate Factors |
| --- | --- | --- | --- |
| Pacific Southwest | March mean temperature | June soil moisture | -0.47 |
| Desert Southwest | July mean temperature | July precipitation | -0.68 |
| Northern Plains | May mean temperature | March PDSI | -0.38 |
| Central Plains | June mean dew point | previous October PDSI | 0.06 |
| Southern Plains | previous October soil moisture | March precipitation | -0.39 |
| Prairie Peninsula | March soil moisture | May mean temperature | -0.46 |
| Ozarks Complex | previous November PDSI | June mean dew point | 0.65 |
| Great Lakes | March soil moisture | January mean temperature | -0.64 |
| Southeast | January soil moisture | May vapor pressure deficit maximum | -0.26 |
| Mid Atlantic | May mean temperature | July precipitation | -0.13 |
| Northeast | previous December soil moisture | May mean temperature | -0.39 |

### Comparing Climate-Informed Retrospective Forecasts vs Benchmarks

In order to quantify the relative forecast skill of the regional climate-informed forecasts, we compared forecast skill against a baseline derived solely from historical WNND caseload. A historical negative binomial baseline model (intercept-only), capturing the distribution of cases historically reported in a given location, has been shown to be a benchmark-to-beat in earlier WNV forecast evaluations^23–25^. We thus fit regional-level historical negative binomial baselines and calculated WNND forecasts at each quantile as described in *Producing Probabilistic Forecasts*.

To assess out-of-sample prediction, we performed a leave-one-out cross validation for both the climate-informed forecasts and historical negative binomial baselines. For example, to create the climate-informed forecast for 2005, we dropped 2005 data from our data subsets (both the climate factor inputs and WNND totals) and re-trained our forecast regression models on the remaining data. Using the resultant models fit to the 2006–2022 (i.e., non-2005) data, we then input the 2005 climate data into the trained model to produce a climate-informed probabilistic retrospective forecast for 2005. This methodology was repeated for all regions and years from 2005–2022. Similarly, we refit a negative binomial baseline to historical data from all years minus the year under consideration; this was repeated for each year and region.

To evaluate the skill of our retrospective forecasts and historical benchmarks, we employed the weighted interval scoring (WIS), a proper scoring metric^42^ that can be considered as a probabilistic version of mean absolute error^43^. WIS is determined directly from quantile forecasts, which made it straightforward to implement here, and is composed of three different error components: dispersion (the specificity of the forecast), and under- and over-prediction (the accuracy of the forecast). These error components can be further analyzed to gather additional information on forecast skill. WIS is zero-bound such that a WIS of zero represents a perfect forecast and higher values of WIS represent worse forecast skill. We calculated WIS as outlined by Bracher et al.^43^, though we first log-transformed the retrospective forecasts and historical benchmarks (adding one before transforming). This adjustment reduces a known bias in WIS where higher valued forecasts lead to higher WIS values^44^. To characterize the uncertainty of regional forecast scores, we bootstrapped WIS of both the retrospective forecasts and historical baselines at the regional-level by resampling yearly WIS with replacement (n = 10,000, sample size = 18 years) across all years and averaging. *P*-values for comparisons between regional retrospective forecast WIS and historical baseline WIS were determined through one-sided pairwise comparisons of the bootstrapped samples. For example, if the retrospective forecast WIS was lower (better skill) than that of the historical baseline for 9,782 of 10,000 pairwise comparisons, the resultant *p*-value for forecast improvement is 0.0218. We used a one-sided comparison test to determine whether the forecast provided improved skill relative to the baseline forecast. Nationally aggregated scores were similarly calculated by averaging across all the regional means determined as described above. This process was repeated with resampling to produce measures of uncertainty (n = 10,000; sample size = 11 regions with 18 years).

A second informative test for evaluating forecast skill is comparison against an ensemble of forecast models. To do so, we utilized the ensemble forecast model created as part of the most recently analyzed CDC WNV Forecasting Challenge (forecasts from the 2022 Challenge are publicly available^25,45^). Ensemble forecasts are generated from a collection of ensemble members, where each member represents forecasts from unique forecast models (often created by different modeling teams) or forecasts from a single forecast model prescribed with varying initial conditions. Ensemble forecasts have frequently demonstrated skill beyond that of the individual ensemble members^46–49^. The ensemble forecast from the 2022 WNV Forecasting Challenge was created by taking the median value of nine individual ensemble member forecasts (the eight forecasting models submitted to the Challenge, as well as a negative binomial historical baseline) at each quantile, such that the ensemble forecast at the 25th percentile for a given county was equal to the median of the 25th percentile forecasts from all ensemble members for that county^25^. To facilitate comparison at the regional level, the ensemble forecast was then aggregated to the regional level using two approaches: 1) directly summing the quantile values across all counties in a given region (i.e., the 50th percentile of the regional forecast is the total of the 50th percentile ensemble forecasts for all counties in the region; a spatially dependent approach; Text S1), and 2) a randomized approach where each individual county-level forecast distribution was sampled and then summed (a spatially independent approach; see Text S2 for complete details). The latter process was repeated 1,000 times and regional-level quantiles were determined from the resulting distributions. WIS was then used to examine the forecast skill of these regional distributions for the 2022 WNV season. We also compared the regional WIS rank of these upscaled forecasts and the climate-informed forecast to ignore region-specific differences of average WIS; a rank of 1 was defined as the best forecast score for a region and 3 was the worst.

### Producing County-Level Forecasts Using a Multi-Level Model

In addition to our regional models, we employed a multi-level model to create a county-level forecast that is conceptually parallel to the regionally determined forecast. In this case, we assumed that the WNND-climate relationship was consistent across each region but with county-specific intercepts distributed around a common mean. We thus regressed county-specific, natural logged WNND counts from 2005–2021 on the same climate variables we determined to be most important to WNND at the regional level (Table 1; Text S1). To facilitate equal comparison with forecast models in the 2022 WNV Forecasting Challenge, we omitted 2022 data from this process. Each regional model was fit as discussed above but including a random county-level intercept (*rstanarm* package in *R*^40,41^). Though not perfectly analogous to the regional forecast, this county-level forecast was methodologically similar and provides a direct comparison with earlier WNV Forecast Challenge forecasts and produces output at the local scale, which is potentially more applicable and actionable.

## Results

### Regional Relationships between Climate and WNV

To guide the development of our climate-informed forecast, we examined the relationship between six climate variables of interest and each month-region combination (see *Investigation of the Influence of Climate on WNND* section; Table 1). Broadly, we found the strongest links between climate factors and annual WNND case counts for measures of soil moisture conditions–whether soil moisture directly or a drought metric like the PDSI–and temperature, though precipitation was regionally important. Notably, we saw multi-regional clustering of a strong relationship between drought and annual WNND caseload over the central third of the U.S. for the 2005–2022 analysis period, extending from the Northern Plains and Great Lakes through to the Southern Plains (Figure 1). These relationships were particularly persistent for the Northern Plains, Prairie Peninsula, and Ozarks Complex regions, where monthly correlations often reached r = −0.6, indicating that increased drought was correlated with higher WNND caseload (note that negative PDSI values are indicative of more intense drought conditions). Similar spatial patterns of relationships emerged for temperature, where warmer winter and spring temperatures were often correlated with higher WNND caseload (Figure 2). These relationships between temperature and annual WNND were often more pronounced in the northern U.S. Heat maps showing the correlations between monthly climate and annual WNND for all regions can be found in the supporting information (Figures S2–S12).

**Figure 1:**
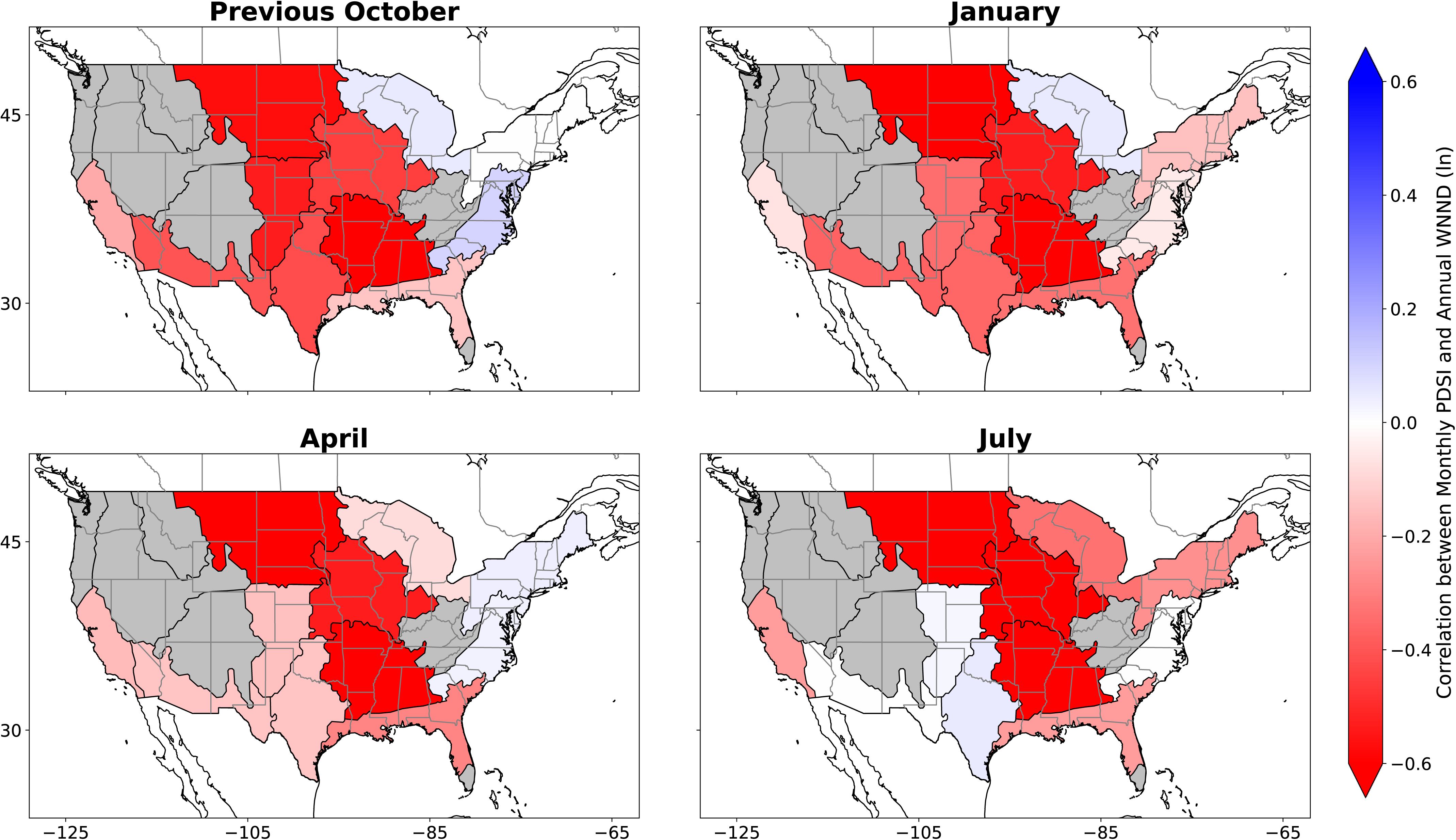
Correlations between Palmer Drought Severity Index and Annual WNND by Region. a) Pearson correlation coefficients between regional natural logged annual WNND caseload and Palmer Drought Severity Index (PDSI) from the preceding October, as well as for the b) January, c) April, and d) July of the concurrent year (red-white-blue fill). Note that PDSI is oriented such that wet conditions are positive so negative correlations denote regions where more intense drought leads to increased WNND caseload. Grey regions were not included in the analysis due to relatively few WNND cases.

**Figure 2:**
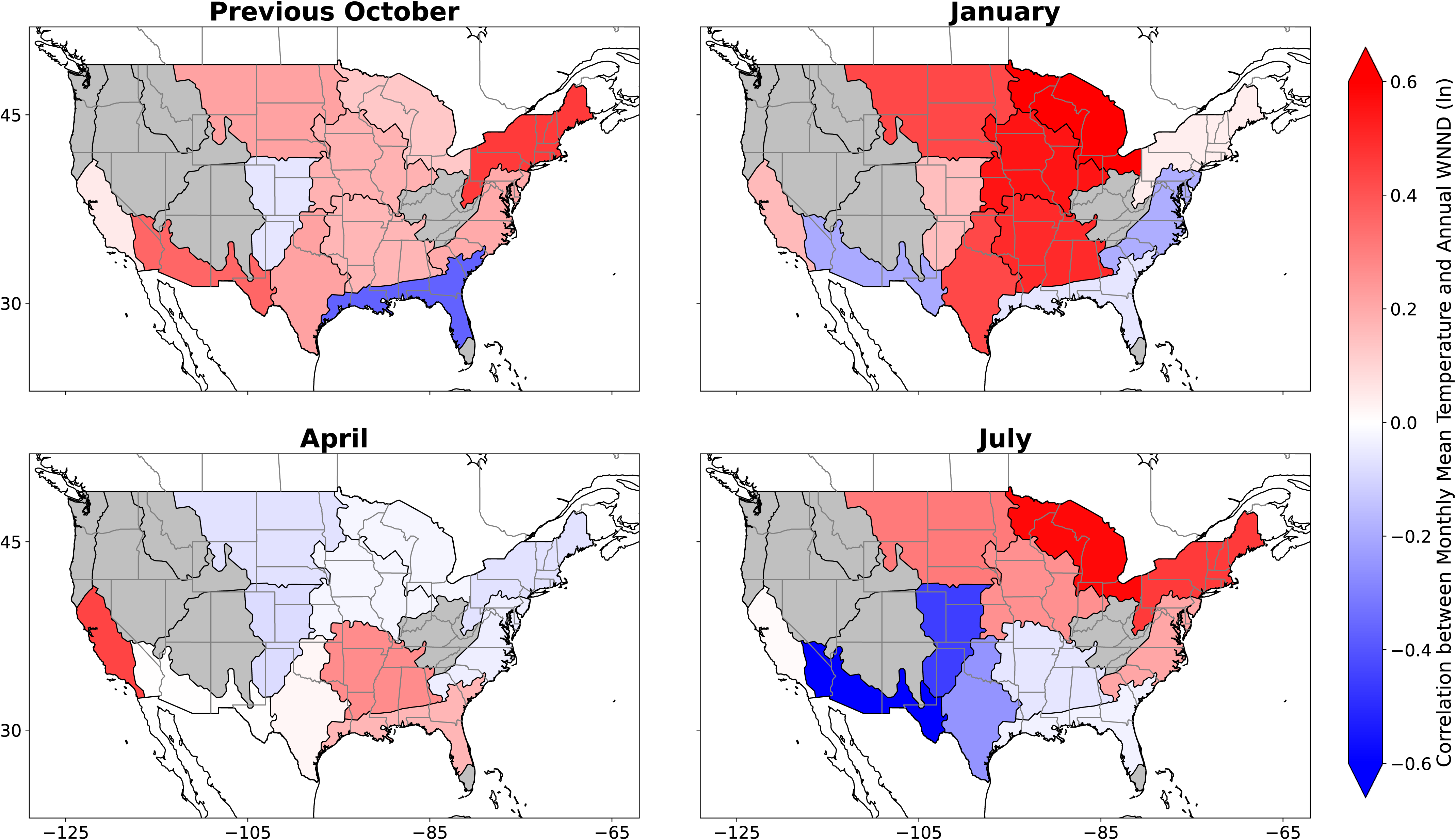
Correlations between Monthly Mean Temperature and Annual WNND by Region. a) Pearson correlation coefficients between regional logged annual WNND caseload and temperature from the preceding October, as well as for the b) January, c) April, and d) July of the concurrent year (red-white-blue fill). Grey regions were not included in the analysis due to relatively few WNND cases.

### Forecast Skill of Climate-Informed Model Compared to Benchmarks

Informed by the climate variable exploratory correlation analysis (Figures 1-2, S2-S12), we constructed our univariate climate-informed WNV forecast model using the climate variable-month combination with the highest absolute correlation for each region (Table 1). We then compared the relative forecast skill of these climate-informed forecasts against benchmarks derived from historical data for 2005–2022. There was a greater than 90% chance of improved forecast skill for the univariate climate-informed forecasts over the historical negative binomial baseline for six regions, as well as a greater than 50% chance of improved forecast skill across all regions (Table S4). Five regions, Ozarks Complex, Northern Plains, Southern Plains, Pacific Southwest, and Prairie Peninsula, all had improved skill (p < 0.05, as determined through bootstrapping and a one-sided comparison test) and had median estimates of 20% or higher improvement compared to the benchmark. Notably, nationally aggregated forecast skill also exceeded the historical negative binomial benchmark with a median forecast skill improvement of 18.1% (95% confidence interval: 3.7–27.0%, p < 0.01). A comparison of WIS components in the univariate model against the historical negative binomial model showed that skill improvements in the univariate model were largely driven by better model calibration (i.e., better accuracy through reduced under- and overprediction). The greatest skill gains were through reduced overprediction, which was accompanied by modest reductions in underprediction and forecast dispersion (Table S2).

We repeated our evaluation of relative retrospective forecast skill for the bivariate climate-informed model. Similarly, the bivariate climate-informed retrospective forecasts demonstrated at least a 90% chance of forecast skill improvements over the historical negative binomial baseline for seven regions, as well as a greater than 50% chance for all 11 regions compared (Table S4). The seven regions of greatest forecast skill improvements showed improvements of 20% or greater (Pacific Southwest, Northern Plains, Central Plains, Southern Plains, Prairie Peninsula, Ozarks Complex, and Mid Atlantic; Figure 3, Table 2). Five of these regions had statistically significant forecast skill improvement (p < 0.05, Northern Plains, Ozarks Complex, Prairie Peninsula, and Mid Atlantic). Nationally aggregated forecast skill for the bivariate model exceeded the historical negative binomial benchmark with an average forecast skill improvement of 23.9% (95% confidence interval: 9.0–32.8%, p < 0.01). An examination of WIS components showed that skill improvements in the bivariate model were largely driven by better model calibration, similar to the findings for the univariate model. While the greatest skill gains were driven by reductions in overprediction, overall skill was further enhanced by modest improvements in both underprediction skill and better forecast dispersion characterizations (Table S3).

**Figure 3:**
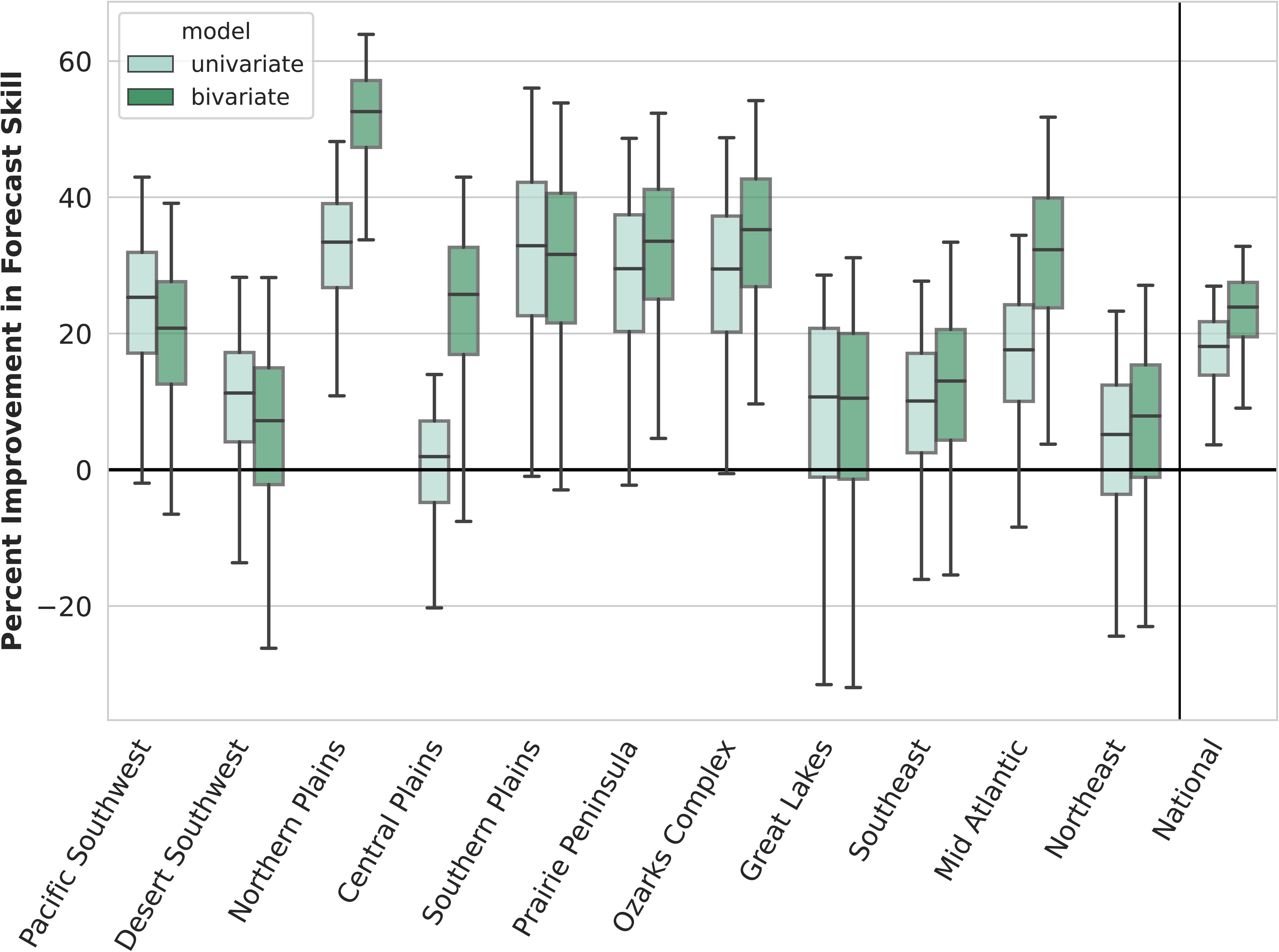
Percent Improvement in Regional Forecast Skill of Climate-Informed Forecast Models Compared to Historical Negative Binomial Benchmark. Box plots of regional and nationally aggregated differences in retrospective forecast skill between univariate (light green) and bivariate (dark green) climate-informed forecast models and historical negative binomial distribution for 2005–2022 as a percentage of historical negative binomial score. Score improvements are calculated as the historical negative binomial weighted interval scores minus the climate-informed forecast model weighted interval scores so that above zero indicate regions where forecast skill is better in the climate-informed forecast model. Box encompasses interquartile range and whiskers depict 95% confidence intervals as determined by bootstrapping. NEON regions are listed from West to East.

**Table 2:** Improvements in Regional Forecast Skill Between Climate-Informed Forecast Models and Historical Negative Binomial Baselines. Regional differences in forecast skill are characterized as the weighted interval score (WIS) of the historical negative binomial WIS minus the univariate or bivariate climate-informed forecast model so that forecast improvements (lower WIS) are shown as positive and are presented as both raw WIS differences and as differences as percentages of the historical negative binomial WIS. The 95% confidence intervals are determined by bootstrapping and shown in brackets as the (2.5^th^, 97.5^th^) percentiles.

| NEON Region | Univariate Forecast |  | Bivariate Forecast |  |
| --- | --- | --- | --- | --- |
|  | Skill Improvement (WIS) | Skill Improvement (%) | Skill Improvement (WIS) | Skill Improvement (%) |
| Pacific Southwest | 0.085 (-0.006, 0.173) | 25.3 (-2, 43) | 0.07 (-0.018, 0.158) | 20.8 (-6.5, 39.1) |
| Desert Southwest | 0.049 (-0.043, 0.15) | 11.3 (-13.7, 28.3) | 0.031 (-0.087, 0.146) | 7.2 (-26.2, 28.2) |
| Northern Plains | 0.196 (0.056, 0.322) | 33.4 (10.9, 48.2) | 0.309 (0.159, 0.463) | 52.6 (33.8, 63.9) |
| Central Plains | 0.008 (-0.061, 0.088) | 1.9 (-20.3, 14) | 0.109 (-0.022, 0.246) | 25.7 (-7.6, 43) |
| Southern Plains | 0.227 (-0.005, 0.477) | 32.9 (-1, 56) | 0.217 (-0.016, 0.471) | 31.6 (-3, 53.9) |
| Prairie Peninsula | 0.128 (-0.008, 0.249) | 29.5 (-2.3, 48.7) | 0.146 (0.016, 0.273) | 33.5 (4.6, 52.3) |
| Ozarks Complex | 0.122 (-0.002, 0.257) | 29.5 (-0.6, 48.8) | 0.147 (0.031, 0.293) | 35.2 (9.7, 54.2) |
| Great Lakes | 0.053 (-0.135, 0.195) | 10.7 (-31.5, 28.6) | 0.051 (-0.127, 0.215) | 10.5 (-32, 31.1) |
| Southeast | 0.041 (-0.05, 0.142) | 10.1 (-16.1, 27.7) | 0.053 (-0.048, 0.169) | 13 (-15.4, 33.4) |
| Mid Atlantic | 0.065 (-0.024, 0.162) | 17.6 (-8.4, 34.4) | 0.119 (0.011, 0.251) | 32.3 (3.8, 51.8) |
| Northeast | 0.02 (-0.081, 0.119) | 5.2 (-24.4, 23.3) | 0.031 (-0.07, 0.139) | 7.9 (-23, 27.1) |
| National Aggregate | 0.092 (0.02, 0.152) | 18.1 (3.7, 27) | 0.119 (0.042, 0.185) | 23.9 (9, 32.8) |

Finally, we compared forecast skill between retrospective forecast skill for the bivariate and univariate climate-informed models (Figure 3; Tables 2, S4). Forecast skill was greater in the bivariate model for 75% or more of paired bootstrapped samples in five regions (Northern Plains, Central Plains, Prairie Peninsula, Ozarks Complex, Mid Atlantic); conversely, forecast skill was greater in the univariate model for 75% or more of bootstrapped samples in two regions (Pacific Southwest, Desert Southwest).

In addition to comparing the climate-informed forecasts against historical benchmarks, we also contrasted the performance of the bivariate model against the ensemble forecast derived from the 2022 CDC WNV Forecasting Challenge. We first upscaled the original county-level ensemble forecast to the regional level utilizing both spatially dependent (aggregating each quantile value across all counties in a given region) and spatially independent methods (aggregating a randomly drawn sample from each county forecast distribution within a given region and then calculating the respective quantiles; see *Comparing Climate-Informed Retrospective Forecasts vs Benchmarks* for more information). We then assessed the climate-informed forecast skill relative to these two regional forecasts for the 2022 WNV season. For this case, the mean of the bivariate climate-informed forecast across regions showed greater skill (WIS = 0.307) than both the spatially independent (WIS = 0.457) and spatially dependent (WIS = 0.522) upscaled ensemble forecasts. When comparing the ordinal ranks of the three forecasts for each region, the climate-informed forecast again demonstrated the most skill (average ranking = 1.5), with the spatially independent forecast in second (average ranking = 2), and the spatially dependent forecast in last (average ranking = 2.5).

Lastly, we examined the skill of the regionally based, multi-level, county-specific forecast against the county-level forecasts from the 2022 Challenge. Using the same bootstrapping method to examine model-specific scores as described above, we found that the climate-informed county-level forecast outperformed 10 of 11 forecasts from the 2022 Challenge (p < 0.05; Table S1), including the historical negative binomial benchmark, and had similar performance to the ensemble forecast. We note that this comparison only included forecasts for counties within the 11 NEON regions included within our analysis and thus did not incorporate all counties included in the 2022 Challenge forecasts.

## Discussion

Here, we created and demonstrated the effectiveness of a regional, climate-informed WNV forecast approach by analyzing retrospective forecast skill. We used regionally determined relationships between climate–using climate data up to July of a given year–and annual WNND to construct both univariate and bivariate climate-informed forecasts of WNND. Our univariate model demonstrated marked improvement in forecast skill compared to a historical benchmark forecast that has performed relatively well in WNV forecasting challenges. Median retrospective forecast skill improved across each of 11 NEON regions, as well as a national aggregate score, with the nationwide forecast and five regions demonstrating significant forecast skill improvements (p < 0.05). Moreover, our bivariate model demonstrated enhanced forecast skill beyond the univariate model, with median retrospective forecast skill improvements for each of the 11 NEON regions compared to the historical negative binomial benchmark and eight of 11 NEON regions compared to the univariate forecast. The additional improvement in nationally aggregated forecast skill resulted in a median 23.9% reduction in weighted interval score over the historical negative binomial benchmark for the bivariate model (95% confidence interval: 9.0–32.8%, p < 0.01). Collectively, these findings demonstrate the value of a regionally derived, climate-informed nationwide WNV forecast approach with substantial improvement over historical benchmarks.

In addition to notable nationwide improvement, our analysis of retrospective forecast skill revealed regional variability in forecast skill changes. The regions which displayed the greatest increase in forecast skill in either the univariate model (> 20% increase in median retrospective forecast skill compared to the historical negative binomial model) or the bivariate model (> 30% increase) were the Northern Plains, Southern Plains, Ozarks Complex, Prairie Peninsula, Pacific Southwest, and Mid Atlantic. Notably, four of these regions are contiguous within the central U.S. and were regional forecasts primarily informed by drought or soil moisture. Combined with our exploratory analysis showing the robust and consistent relationship between PDSI and WNND in this portion of the country (Figure 1), there is strong evidence that drought is a dominant climate factor on WNV in this area and that incorporating drought into WNV forecast efforts is important for enhancing forecast skill. In comparison, the regions with the least improvement in forecast skill across both models were the Desert Southwest, Great Lakes, Northeast, and Southeast regions. While additional analysis is needed to determine if there are causal factors of lessened forecast improvements in these areas, we note that the Northeast and Southeast regions have the fewest total WNND cases of the regions we examined (Figure S1) and the Desert Southwest experienced a dramatic outbreak in 2021 (37% of cases occurred in this year), which may skew the statistical regressions underlying model development.

As mentioned above, there were also differences in which climate factors were used in our models. While our methodology allowed for regional differences in incorporated climate factors, some broad trends did emerge. Notably, antecedent soil moisture, characterized either through PDSI or 50-mm soil moisture, was the primary climate factor for six of the 11 regions under consideration–including a five region cluster covering the central third of the U.S.–and was the secondary climate factor for three additional regions. Temperature was incorporated into our models in a total of seven regions, including as the primary climate factor for four of the remaining five regions. Three additional climate factors–precipitation (three times), dew point (two), and maximum vapor pressure deficit (one)–were used, though less consistently. Of note, while the months of the incorporated drought data ranged widely throughout the year, including the previous fall for several regions, the months used for the remaining climate factors were closer to the summer WNV peak (all but one in March–July). This may be due to the greater persistence of drought throughout the year (see Figures S6-S8 for examples of the consistency in relationship between PDSI and WNND), compared to more transient climatic conditions like temperature and precipitation, or attributable to the precise biological or ecological mechanisms in which these factors affect WNV transmission and its vectors.

While both the univariate and bivariate climate-informed models demonstrated increased forecast skill compared to historical negative binomial benchmarks, there were some differences between the two models. Notably, the bivariate model outperformed the univariate model in retrospective forecast performance for greater than 75% of bootstrapped samples over a grouping of regions in the central U.S.–the Northern Plains (28.8% median improvement in bivariate compared to univariate), Central Plains (23.7%), Ozarks Complex (9.0%), Prairie Peninsula (6.1%), and the Mid Atlantic (18.1%)–while the univariate model consistently outperformed the bivariate model in the Pacific Southwest (5.8%) and Desert Southwest (4.4%) regions. These findings, in addition to the bivariate national aggregate score outperforming the univariate national aggregate score in nearly 99% of samples (7.3% median improvement), provide evidence that adding additional climate information to regional forecasts generally boosted forecast skill, though this result was not universal as three of 11 regions did not show bivariate model superiority. Regional discrepancies in the additional value provided by incorporating a second climate factor may be tied to differences in the covariance of the input climate factors or by the complementing or offsetting influences of multiple climate variables.

In order to determine how the regional-level forecast approach improves retrospective forecast skill, we dissected the components of the WIS: forecast dispersion, overprediction, and underprediction. This allowed us to identify the changes in forecast components providing the greatest boost in forecast skill. Both of our climate-informed models show the majority of total skill increases were driven by reductions in overprediction, with improvements in overprediction generating 71% and 64% of overall skill changes in the univariate and bivariate models, respectively. Both models also show small skill gains through reductions in underprediction (15% and 19% of total univariate and bivariate skill improvements, respectively), and better characterization of forecast dispersion (15% and 18% of total univariate and bivariate skill improvements, respectively). These findings suggest that the improvements in forecast skill at the regional level, primarily driven by reductions in overprediction, are due to better constrained forecasts that effectively capture reductions in the likelihood of high caseload outbreak events. This improvement may provide public health utility through better characterization of the likelihood of relatively high- or low-risk WNV seasons.

Our findings also show the importance of spatiotemporal scale in being able to identify the relationships between WNND and climate or other environmental factors. Given the relative sparseness of WNND at the level of the individual county (99% of counties average one or fewer cases of WNND per year), attempts to determine the underlying influences of climate or other environmental factors on WNND caseload at the county level across the U.S. have proved challenging. Scaling limitations such as these lead to the problem of scale being described as the “central problem in ecology”^50^. We combat these inherent constraints by aggregating our data to a regional scale based on eco-climatic zones and performing our analysis at that higher level. This aggregation leads to higher annual WNND counts and, accordingly, a greater interannual statistical signal to compare against climate factors previously shown to be biologically or ecologically relevant to WNV or its vectors^26,51^. However, selecting the appropriate division of regions on which to perform this analysis leaves many options, and there exists a wide array of regional divisions to choose between. While regions are often derived from administrative boundaries (i.e., U.S. states or counties; administrative level one or two, respectively), we center our analysis on regions that are ecologically and climatically derived (i.e., NEON regions) and found evidence of benefits to this approach. This broader notion of using ecologically meaningful regions is supported by recent work on WNV^51^ as, while the problem posed by WNV is a public health one, understanding the underlying variability in an effort to inform public health responses is fundamentally an ecological problem.

Beyond the inescapable nuanced trade-offs within our regional methodology–including both the selection of the regions, as well as lack of specificity produced by regionally based approaches–we recognize several potential limitations of our methodology. We focused our analysis on the influence of monthly climate conditions (e.g., March temperature) on annual WNND cases in an effort to identify the time periods most critical to annual WNV risk. It is possible that additional forecast improvements could be made by focusing on either longer, seasonal (e.g., March–May) or shorter, weekly (e.g., April 15–21) time windows. It is also possible that constraining our analysis to calendar months may inadvertently divide critical times and dampen the resulting empirical relationships we characterize. For example, if WNV in a particular region is critically dependent on temperature across the last two weeks of March and first two weeks of April, or if the timing shifts between years, focusing on either March or April temperatures will result in a reduced statistical signal compared to what is theoretically possible. We note that our analysis allowed for the selection of time windows as late into the calendar year as July, with July climate variables selected in two regions. While these windows do provide insight into what is expected during the peak of the upcoming WNV season, being able to provide forecasts based on climatic conditions earlier in the year would provide valuable additional lead time. Additionally, our focus on forecasts of annual WNND leaves an opportunity for future research to examine shifts in WNV seasonal timing that could provide valuable insight into an additional and useful dimension of a WNV forecast: will the upcoming WNV season be earlier or later than usual?

We also acknowledge that our exploratory methodology to identify climate variables of importance may result in overfitting of our forecast models. While we did select the climate variables that provide the basis of our WNV forecast based on the strongest correlations, we focused additional analyses on out-of-sample predictions by 1) producing the evaluative retrospective forecasts using a leave-one-out methodology, 2) imposing restrictions on the climate variables we selected for the bivariate model (i.e., the variables must be from different categories–near-surface air conditions, antecedent soil moisture conditions, and precipitation; this limited r-squared coefficients of determination to < 0.25 for all but three regions), and 3) focusing our exploratory analysis on climate inputs shown to be biologically or ecologically important. Conversely, it may be possible to further boost forecast abilities through a more rigorous examination of how potential input climate factors impact forecast skill beyond correlating monthly climate and WNND, including an examination of climate factor covariance and interactions for the bivariate forecast model. Additionally, our analysis does not account for any positive or negative temporal autocorrelation that may be caused by, for example, persistence of WNV or WNV immunity in host avian and human populations from one year to the next^9^.

While our regional approach demonstrated skill through retrospective forecasts of WNV, translating these forecasts to informative prospective forecasts at the more actionable county or state scales remains challenging. To begin, our retrospective forecasts were designed using historical data, but creating real-time forecasts will require consideration of climate data publication latency. Notably, while the PRISM, gridMET, and NLDAS products are all available in near real-time (i.e., within several days), this may constrain forecast availability such that forecasts requiring July data would not be available until a few days after the end of July. Additionally, PRISM climate data is considered provisional until six months in the future to allow for delays in station reporting^31^. As noted above, our analysis allowed for the selection of time windows as late into the calendar year as July. While this remains in advance of the majority of WNV burden in a typical season, it is not conducive to providing extensive forecast lead times before the typical August–September peak. Forecast models could be constructed around earlier climate observations to provide additional lead time, though this may result in reduced forecast accuracy compared to what is shown here. Producing prospective forecasts available at specific planning horizons will require balancing the needs and constraints of decision-maker needs, data availability, and forecast accuracy (see Keyel et al., 2021^52^ and Hii et al., 2012^53^ for additional discussion on this topic).

It may be possible to extend the lead time of this, and similar, climate-informed disease models through the incorporation of daily to seasonal weather and climate forecasts. The use of short-term (i.e., one to two weeks) weather forecasts may allow for the extension of disease forecast lead times with relatively high accuracy, while subseasonal-to-seasonal (i.e., one to six months) near-term climate forecasts may enable more extended lead times for particular forecasts of opportunity (i.e., interannual variability in WNND caseload in Arizona may coincide with a potentially more predictable climate phenomenon, the North American Monsoon^54,55^). More research is needed to characterize the potential benefits and limitations of this unexplored approach to disease forecasting. It would also be ideal to improve forecasting across scales as more actionable WNV forecasts would likely have the greatest benefit at the local scales of public health and vector control agency decision-makers. While our approach to generating a downscaled, county-level forecast based on regional climate-WNV relationships produced statistically significant improvements compared to the historical negative binomial benchmark, it provided only a marginal 2.3% increase in county-level forecast skill. Future work should build upon our findings by aiming to produce a more localized, and thus actionable, forecast derived from our regional approach.

### Summary

Using retrospective forecasts, we demonstrated the improved skill of a regionally defined, climate-informed forecast of WNV for the continental U.S. We developed this forecast approach by first examining the associations between a wide array of biologically meaningful climate factors and WNND caseload in the most affected regions of the U.S.; the most influential climate factors for each region were then incorporated into regional forecast models. Comparing the forecast skill of our climate-informed models against historically derived benchmarks from 2005 through 2022 showed substantial improvements in nationwide forecast skill with a median reduction in nationally aggregated weighted interval score of 18% and 24% for our univariate and bivariate climate-informed WNV forecast models, respectively. An additional analysis showed improvements of these forecasts over regionally aggregated county-level ensemble forecasts from the 2022 WNV Forecasting Challenge. Moreover, a climate-informed county-level retrospective forecast showed greater forecast skill than all forecasts in the 2022 WNV Forecasting Challenge, including both the ensemble and the historical negative binomial benchmark, though improvement over the ensemble forecast was not statistically significant.

To our knowledge, this represents the first nationwide WNV forecast model with demonstrated improvements over simple historically derived, distribution-based baseline forecasts^23,25^. The findings shown here, in particular the methodological advancements facilitated by a regional focus, represent steps toward actionable U.S. WNV forecasts and a foundation to build toward effective informational products. Additional research is needed to build useful prospective forecasts from the regional approach described here. One such step would be to determine how to leverage gains from regionally derived forecasts to improve smaller-scale forecasts (e.g., county-level) that may be more conducive to decision-maker needs. A further enhancement would determine the potential usability of weather and climate forecasts as inputs into climate-sensitive disease forecasts^56,57^. While these inputs would be less precise than incorporating observed climatic conditions, they may also lead to valuable additional lead time. Finally, we suggest examining links between the timing of the WNV season and local climate conditions to further characterize the influence of climate on WNV burden.

## Data Availability

Data sources for both U.S. West Nile virus case counts and climate data are publicly available at the corresponding links:
West Nile virus neuroinvasive disease counts (https://www.cdc.gov/west-nile-virus/data-maps/historic-data.html), PRISM (https://prism.oregonstate.edu/), gridMET (https://www.climatologylab.org/gridmet.html), NLDAS (https://ldas.gsfc.nasa.gov/nldas).
Finally, code developed by the authors for the data analysis and visualization is openly available at https://github.com/ryandharp/regionally_determined_climate-informed_wnv_forecast and will be preserved in a public repository (e.g., Zenodo) for posterity upon publication.

https://prism.oregonstate.edu/

https://www.climatologylab.org/gridmet.html

https://ldas.gsfc.nasa.gov/nldas

## Acknowledgments

We thank all who contributed to the CDC ArboNET WNV data set, including data collection, reporting, and cleaning. We also thank the PRISM Climate Group at Oregon State University; the Climatology Lab at the University of California, Merced; and the NLDAS project team for producing the publicly available climate data in the PRISM, gridMET, and NLDAS data sets. We also express our gratitude to many others who have provided their support and insight and support on this work, especially Randall J. Nett, J. Erin Staples, C. Ben Beard, Lyle R. Peterson, and Juli Trtanj.

RDH acknowledges the NOAA-Climate Adaptation and Mitigation Program for the support administered by UCAR’s Cooperative Programs for the Advancement of Earth System Science (CPAESS) under award NA21OAR4310473. SGB and BWG were supported by the NOAA Cooperative Agreement NA22OAR4320151 for the Cooperative Institute for Earth System Research and Data Science (CIESRDS). SGB and BWG were also partially supported by the National Science Foundation (NSF) and its Directorate for Geosciences Climate Change Impacts on Human Health (C2H2) via NSF Award 2432999. None of the funding bodies had a role in the design of the study and collection, analysis, and interpretation of data and in writing the manuscript. The findings and conclusions in this report are those of the authors and do not necessarily represent the views of the Centers for Disease Control and Prevention, the Department of Health and Human Services, the National Oceanic and Atmospheric Administration, or the Department of Commerce.

Data sources for both U.S. West Nile virus case counts and climate data are publicly available at the corresponding links:

- West Nile virus neuroinvasive disease counts (https://www.cdc.gov/west-nile-virus/data-maps/historic-data.html)
- PRISM (https://prism.oregonstate.edu/)
- gridMET (https://www.climatologylab.org/gridmet.html)
- NLDAS (https://ldas.gsfc.nasa.gov/nldas)

Finally, code developed by the authors for the data analysis and visualization is openly available at https://github.com/ryandharp/regionally_determined_climate-informed_wnv_forecast and will be preserved in a public repository (e.g., Zenodo) for posterity upon publication.

## References

1. Nash, D., Mostashari, F., Fine, A., Miller, J., O’leary, D., Murray, K.,… & Layton, M. (2001). The outbreak of West Nile virus infection in the New York City area in 1999. New England Journal of Medicine, 344(24), 1807–1814.

2. The U.S. Department of Health and Human Services and the U.S. Centers for Disease Control and Prevention. The National Public Health Strategy to Prevent and Control Vector-Borne Diseases in People. 2024. https://www.cdc.gov/vector-borne-diseases/php/data-research/national-strategy/index.html. Accessed 12 Sept 2024.

3. Zou S, Foster GA, Dodd RY, Petersen LR, Stramer SL. West Nile fever characteristics among viremic persons identified through blood donor screening. The Journal of infectious diseases. 2010 Nov 1;202(9):1354–61.

4. Mostashari F, Bunning ML, Kitsutani PT, Singer DA, Nash D, Cooper MJ, et al. Epidemic West Nile encephalitis, New York, 1999: results of a household-based seroepidemiological survey. The Lancet. 2001 Jul 28;358(9278):261–4.

5. McDonald E. Surveillance for West Nile virus disease—United States, 2009–2018. MMWR. Surveillance Summaries. 2021;70.

6. Hughes JM, Wilson ME, Sejvar JJ. The long-term outcomes of human West Nile virus infection. Clinical infectious diseases. 2007 Jun 15;44(12):1617–24.

7. CDC. (2024, May 17). Data and Maps for West Nile. West Nile Virus. https://www.cdc.gov/west-nile-virus/data-maps/index.html

8. Gage, K. L., Burkot, T. R., Eisen, R. J., & Hayes, E. B. (2008). Climate and vectorborne diseases. American journal of preventive medicine, 35(5), 436–450.

9. Paull, S. H., Horton, D. E., Ashfaq, M., Rastogi, D., Kramer, L. D., Diffenbaugh, N. S., & Kilpatrick, A. M. (2017). Drought and immunity determine the intensity of West Nile virus epidemics and climate change impacts. Proceedings of the Royal Society B: Biological Sciences, 284(1848), 20162078.

10. Shocket, M. S., Verwillow, A. B., Numazu, M. G., Slamani, H., Cohen, J. M., El Moustaid, F., Rohr, J., Johnson, L. R., & Mordecai, E. A. (2020). Transmission of West Nile and five other temperate mosquito-borne viruses peaks at temperatures between 23 C and 26 C. Elife, 9, e58511.

11. Reisen, W. K., Fang, Y., & Martinez, V. M. (2014). Effects of temperature on the transmission of West Nile virus by Culex tarsalis (Diptera: Culicidae). Journal of medical entomology, 43(2), 309–317.

12. Beard, C. B., Eisen, R. J., Barker, C. M., Garofalo, J. F., Hahn, M., Hayden, M.,… & Schramm, P. J. (2016). Ch. 5: Vector-borne diseases (pp. 129–156). US Global Change Research Program, Washington, DC.

13. Chuang, T. W., Hildreth, M. B., Vanroekel, D. L., & Wimberly, M. C. (2011). Weather and land cover influences on mosquito populations in Sioux Falls, South Dakota. Journal of medical entomology, 48(3), 669–679.

14. Gardner, A. M., Hamer, G. L., Hines, A. M., Newman, C. M., Walker, E. D., & Ruiz, M. O. (2012). Weather variability affects abundance of larval Culex (Diptera: Culicidae) in storm water catch basins in suburban Chicago. Journal of medical entomology, 49(2), 270–276.

15. Koenraadt, C. J. M., & Harrington, L. C. (2008). Flushing effect of rain on container-inhabiting mosquitoes Aedes aegypti and Culex pipiens (Diptera: Culicidae). Journal of medical entomology, 45(1), 28–35.

16. Hahn, M. B., Monaghan, A. J., Hayden, M. H., Eisen, R. J., Delorey, M. J., Lindsey, N. P., Nasci, R. S., & Fischer, M. (2015). Meteorological conditions associated with increased incidence of West Nile virus disease in the United States, 2004–2012. The American journal of tropical medicine and hygiene, 92(5), 1013.

17. Wimberly, M. C., Lamsal, A., Giacomo, P., & Chuang, T. W. (2014). Regional variation of climatic influences on West Nile virus outbreaks in the United States. The American journal of tropical medicine and hygiene, 91(4), 677.

18. Chase, J. M., & Knight, T. M. (2003). Drought-induced mosquito outbreaks in wetlands. Ecology Letters, 6(11), 1017–1024.

19. Russell, M. C., Herzog, C. M., Gajewski, Z., Ramsay, C., El Moustaid, F., Evans, M. V., Desai, T., Gottdenker, N. L., Hermann, S. L., Power, A. G., & McCall, A. C. (2022). Both consumptive and non-consumptive effects of predators impact mosquito populations and have implications for disease transmission. Elife, 11, e71503.

20. Brown, J. J., Pascual, M., Wimberly, M. C., Johnson, L. R., & Murdock, C. C. (2023). Humidity–The overlooked variable in the thermal biology of mosquito-borne disease. Ecology letters, 26(7), 1029–1049.

21. Wimberly, M. C., Davis, J. K., Hildreth, M. B., & Clayton, J. L. (2022). Integrated forecasts based on public health surveillance and meteorological data predict West Nile virus in a high-risk region of North America. Environmental Health Perspectives, 130(8), 087006.

22. Fyie, L. R., Gardiner, M. M., & Meuti, M. E. (2021). Artificial light at night alters the seasonal responses of biting mosquitoes. Journal of Insect Physiology, 129, 104194.

23. Holcomb, K. M., Mathis, S., Staples, J. E., Fischer, M., Barker, C. M., Beard, C. B., Nett, R. J., Keyel, A. C., Marcantonio, M., Childs, M. L., Gorris, M. E., Rochlin, I., Hamins-Puértolas, M., Ray, E. L., Uelmen, J. A., DeFelice, N., Freedman, A. S., Hollingsworth, B. D., Das, P., Osthus, D., Humphreys, J. M., Nova, N., Mordecai, E. A., Cohnstaedt, L. W., Kirk, D., Kramer, L. D., Harris, M. J., Kain, M. P., Reed, E. M. X., & Johansson, M. A. (2023a). Evaluation of an open forecasting challenge to assess skill of West Nile virus neuroinvasive disease prediction. Parasites & Vectors, 16(1), 11.

24. Holcomb, K. M., Staples, J. E., Nett, R. J., Beard, C. B., Petersen, L. R., Benjamin, S. G., Green, B. W., Jones, H., & Johansson, M. A. (2023b). Multi-model prediction of West Nile virus neuroinvasive disease with machine learning for identification of important regional climatic drivers. GeoHealth, 7(11), e2023GH000906.

25. Harp, R. D., Holcomb, K. M., Retkute, R., Prusokiene, A., Prusokas, A., Ertem, Z., Ajelli, M., Kummer, A. G., Litvinova, M., Merler, S., Pastore y Piontti, A., Poletti, P., Vespignani, A., Wilke, A. B. B., Zardini, A., Smith, K. H., Armstrong, P., DeFelice, N., Keyel, A., Shepard, J., Smith, R., Tyre, A., Humphreys, J., Cohnstaedt, L., Hosseini, S., Scoglio, C., Gorris, M., Barnard, M., Moser, S. K., Spencer, J., McCarter, M. S. J., Lee, C., Nolan, M. S., Barker, C. M., Staples, J. E., Nett, R. J., & Johansson, M. A. (in press). Evaluation of the 2022 West Nile Virus Forecasting Challenge, United States. Parasites & Vectors.

26. Uelmen, J. A., Irwin, P., Bartlett, D., Brown, W., Karki, S., Ruiz, M. O. H.,… & Smith, R. L. (2021). Effects of scale on modeling West Nile virus disease risk. The American Journal of Tropical Medicine and Hygiene, 104(1), 151.

27. DeFelice NB, Little E, Campbell SR, Shaman J. Ensemble forecast of human West Nile virus cases and mosquito infection rates. Nature Communications. 2017 Feb 24;8(1):14592.

28. Little E, Campbell SR, Shaman J. Development and validation of a climate-based ensemble prediction model for West Nile Virus infection rates in Culex mosquitoes, Suffolk County, New York. Parasites & Vectors. 2016 Dec;9:1–3.

29. Danforth ME, Snyder RE, Lonstrup ET, Barker CM, Kramer VL. Evaluation of the effectiveness of the California mosquito-borne virus surveillance & response plan, 2009–2018. PLoS neglected tropical diseases. 2022 May 9;16(5):e0010375.

30. Ward MJ, Sorek-Hamer M, Henke JA, Little E, Patel A, Shaman J, et al. A spatially resolved and environmentally informed forecast model of West Nile virus in Coachella Valley, California. GeoHealth. 2023 Dec;7(12):e2023GH000855.

31. PRISM Climate Group, & Oregon State University. (2023). Monthly mean temperature, minimum temperature, and total precipitation datasets 1999-2021 [Dataset]. Retrieved from https://prism.oregonstate.edu.

32. Abatzoglou, J. T. (2013), Development of gridded surface meteorological data for ecological applications and modelling. Int. J. Climatol., 33: 121–131.

33. Xia, Y., Mitchell, K., Ek, M., Sheffield, J., Cosgrove, B., Wood, E., Luo, L., Alonge, C., Wei, H., Meng, J., Livneh, B., Lettenmaier, D., Koren, V., Duan, Q., Mo, K., Fan, Y., & Mocko, D. (2012). Continental-scale water and energy flux analysis and validation for the North American Land Data Assimilation System project phase 2 (NLDAS-2): 1. Intercomparison and application of model products. Journal of Geophysical Research: Atmospheres, 117(D3).

34. National Ecological Observatory Network. (n.d.). Spatial and temporal design. NEON science. Retrieved from https://www.neonscience.org/about/overview/design.

35. U.S. Census Bureau (2024). TIGER/Line Shapefiles. Census.gov. https://www.census.gov/geographies/mapping-files/time-series/geo/tiger-line-file.2022.html#list-tab-790442341

36. U.S. Census Bureau (2024). Centers of Population. Census.gov. https://www.census.gov/geographies/reference-files/time-series/geo/centers-population.html

37. NSF NEON (n.d.). Spatial and Temporal Design. www.neonscience.org. Retrieved June 5, 2024, from https://www.neonscience.org/about/overview/design

38. Gorris, M. E., Bartlow, A. W., Temple, S. D., Romero-Alvarez, D., Shutt, D. P., Fair, J. M., Kaufeld, K. A., Del Valle, S. Y., & Manore, C. A. (2021). Updated distribution maps of predominant Culex mosquitoes across the Americas. Parasites & Vectors, 14, 1–13.

39. Rochlin, I., Faraji, A., Healy, K., & Andreadis, T. G. (2019). West Nile virus mosquito vectors in North America. Journal of Medical Entomology, 56(6), 1475–1490.

40. Goodrich, B., Gabry J., Ali I., & Brilleman, S. (2023). rstanarm: Bayesian applied regression modeling via Stan. R package version 2.26.1 https://mc-stan.org/rstanarm.

41. R Core Team (2023). R: A Language and Environment for Statistical Computing. R Foundation for Statistical Computing, Vienna, Austria. https://www.R-project.org/.

42. Gneiting T and AE Raftery. (2007) Strictly proper scoring rules, prediction, and estimation. Journal of the American Statistical Association. 102(477):359–378. Available at: https://www.stat.washington.edu/raftery/Research/PDF/Gneiting2007jasa.pdf.

43. Bracher, J., Ray, E. L., Gneiting, T., & Reich, N. G. (2021). Evaluating epidemic forecasts in an interval format. PLoS computational biology, 17(2), e1008618.

44. Bosse, N. I., Abbott, S., Cori, A., van Leeuwen, E., Bracher, J., & Funk, S. (2023). Scoring epidemiological forecasts on transformed scales. PLoS Computational Biology, 19(8), e1011393.

45. Github. Data and forecast submission repository for the 2022 CDC West Nile virus Forecasting Challenge. https://github.com/cdcepi/WNV-forecast-data-2022. Accessed 12 Sept 2024.

46. Cramer, E. Y., Ray, E. L., Lopez, V. K., Bracher, J., Brennen, A., Castro Rivadeneira, A. J.,… & Georgescu, A. (2022). Evaluation of individual and ensemble probabilistic forecasts of COVID-19 mortality in the United States. Proceedings of the National Academy of Sciences, 119(15), e2113561119.

47. Johansson, M. A., Apfeldorf, K. M., Dobson, S., Devita, J., Buczak, A. L., Baugher, B.,… & Chretien, J. P. (2019). An open challenge to advance probabilistic forecasting for dengue epidemics. Proceedings of the National Academy of Sciences, 116(48), 24268–24274.

48. Reich, N. G., McGowan, C. J., Yamana, T. K., Tushar, A., Ray, E. L., Osthus, D., Kandula, S., Brooks, L. C., Crawford-Crudell, W., Gibson, G. C., Moore, E., Silva, R., Biggerstaff, M., Johansson, M. A., Rosenfeld, R., & Shaman, J. (2019). Accuracy of real-time multi-model ensemble forecasts for seasonal influenza in the US. PLoS computational biology, 15(11), e1007486.

49. McGowan, C. J., Biggerstaff, M., Johansson, M., Apfeldorf, K. M., Ben-Nun, M., Brooks, L., Convertino, M., Erraguntla, M., Farrow, D. C., Freeze, J., Ghosh, S., Hyun, S., Kandula, S., Lega, J., Liu, Y., Michaud, N., Morita, H., Niemi, J., Ramakrishnan, N., Ray, E. L., Reich, N. G., Riley, P., Shaman, J., Tibshirani, R., Vespinani, A., Zhang, Q., Reed, C., & The Influenza Forecasting Working Group (2019). Collaborative efforts to forecast seasonal influenza in the United States, 2015–2016. Scientific reports, 9(1), 683.

50. Levin, S. A. (1992). The problem of pattern and scale in ecology: the Robert H. MacArthur award lecture. Ecology, 73(6), 1943–1967.

51. Moser, S. K., Spencer, J. A., Barnard, M., Hyman, J. M., Manore, C. A., & Gorris, M. E. (2024). Exploring climate-disease connections in geopolitical versus ecological regions: The case of West Nile virus in the United States. GeoHealth, 8(6), e2024GH001024.

52. Keyel, A. C., Gorris, M. E., Rochlin, I., Uelmen, J. A., Chaves, L. F., Hamer, G. L.,… & Smith, R. L. (2021). A proposed framework for the development and qualitative evaluation of West Nile virus models and their application to local public health decision-making. PLoS neglected tropical diseases, 15(9), e0009653.

53. Hii, Y. L., Rocklöv, J., Wall, S., Ng, L. C., Tang, C. S., & Ng, N. (2012). Optimal lead time for dengue forecast. PLoS neglected tropical diseases, 6(10): e1848.

54. Holcomb, K. M. (2022). Worst-ever US West Nile virus outbreak potentially linked to a wetter-than-average 2021 Southwest monsoon. Silver Spring (MD): Climate.gov.

55. Prein, A. F., Towler, E., Ge, M., Llewellyn, D., Baker, S., Tighi, S., & Barrett, L. (2022). Sub-seasonal predictability of North American monsoon precipitation. Geophysical Research Letters, 49(9), e2021GL095602.

56. Merryfield, W. J., Baehr, J., Batté, L., Becker, E. J., Butler, A. H., Coelho, C. A.,… & Yeager, S. (2020). Current and emerging developments in subseasonal to decadal prediction. Bulletin of the American Meteorological Society, 101(6), E869–E896.

57. White, C. J., Domeisen, D. I., Acharya, N., Adefisan, E. A., Anderson, M. L., Aura, S.,… & Wilson, R. G. (2022). Advances in the application and utility of subseasonal-to-seasonal predictions. Bulletin of the American Meteorological Society, 103(6), E1448–E1472.

