## Supplemental Material for "A Regionally Determined Climate-Informed West Nile Virus Forecast Technique"

***Text S1: Descriptions of Model Development***

We used a generalized linear Bayesian regression model to produce probabilistic climate-informed regional-level forecasts of annual WNND cases. This was performed for both univariate and bivariate climate-informed forecasts to determine the influence of X_i_ regional climate variables on annual WNND cases. These regressions were fit using the stan_glm.nb function in the *rstanarm* package in *R* (Goodrich et al., 2023; R Core Team, 2023) and are of the following form:

$cases \sim Negative Binomial(\mu_{i},\phi)$

$\mu_{i}=\beta_{0}+\beta_{1}X_{1}$(univariate form) or

$\mu_{i}=\beta_{0}+\beta_{1}X_{1}+\beta_{2}X_{2}$(bivariate form)

where μ is the mean, 𝜙 is the dispersion, β_0_ is the intercept, and β_1_ and β_2_ are fit regression coefficients for the selected climate covariate. We ran four chains for each regression with a burn-in of 500 samples, then collected an additional 500 for each chain, resulting in 2,000 total samples across the four chains.

We used a similar generalized linear mixed-effects regression model in a Bayesian framework to produce probabilistic climate-informed county-level forecasts of annual WNND cases driven by regional-level climate conditions. This was also performed for both univariate and bivariate climate-informed forecasts to determine the influence of X_i_ regional climate variables on county-level annual WNND cases, with county as a random effect. These regressions were fit using the stan_glmer.nb function in the *rstanarm* package in *R* (Goodrich et al., 2023; R Core Team, 2023) and had the following form:

$cases \sim Negative Binomial(\mu_{i},\phi)$

$\mu_{i}=\beta_{0}+\beta_{1}X_{1}+(1 | county)$ (univariate form) or

$\mu_{i}=\beta_{0}+\beta_{1}X_{1}+\beta_{2}X_{2}+(1 | county)$(bivariate form)

where μ is the mean, $\phi$ is the dispersion, β_0_ is the intercept, and β_1_ and β_2_ are regression coefficients. We ran four chains for each regression with a burn-in of 500 samples, then collected an additional 500 for each chain, resulting in 2,000 total samples across the four chains.

***Text S2: Description of Regional Aggregation of Ensemble Forecasts***

We used two approaches to produce regional benchmark forecasts from county-level ensemble forecasts from the 2022 West Nile Virus Forecasting Challenge. First, we performed a spatially dependent approach where the *q*-quantile forecast for a given region is equal to the sum of the *q*-quantile forecast values from all counties within that region:

${forecast}_{region, q} = \sum_{i=1}^{c} {forecast}_{i, q}$

where c is the set of all counties in a region and *q* is the set of 23 quantiles in {0.01, 0.025, 0.05, 0.10, …, 0.90, 0.95, 0.975, 0.99}.

We also applied a spatially independent approach where the *q*-quantile forecast for a given region was generated from a distribution of the sum of counts sampled randomly across each county in each region (n = 1,000). Each bootstrapped regional forecast is generated by adding one random sample from each c county forecast within the region. We denote forecast*_region_ as a sample regional forecast determined from a single bootstrapped sample across all c counties in the region:

${forecast*}_{region}=\sum_{i=1}^{c} N_{i}$

such that N_i_ is a random sample from the county-specific distribution of case forecasts. The *q*-quantile forecast using n = 1,000 bootstrapped samples for that region is then:

${forecast}_{region, q} = Q\left( {forecast*}_{region, 1...n}, q \right)$

where $Q$ is the quantile at $q$ $in \{0.01, 0.025, 0.05, 0.10, .. , 0.90, 0.95, 0.975, 0.99\}$.

***Table S1: Comparison of Multi-Level County-Specific Climate-Informed WNV Forecast with 2022 CDC WNV Forecasting Challenge Forecast Models.***

| **Comparison Model** | **Improvement in Mean Score (%)** | **p-value** |
| --- | --- | --- |
| ensemble | 1.0 | n.s. |
| historical negative binomial | 2.3 | 0.011 |
| hybrid-hybrid | 3.8 | 0.001 |
| MSSM-WED | 5.6 | <0.001 |
| LANL-NBandP | 8.8 | <0.001 |
| Datart-PoissonFE | 17.1 | <0.001 |
| AMbeRland-RandomForest_anomaly | 28.9 | <0.001 |
| FINforWN-MCMaWN | 33.3 | <0.001 |
| CDC-NaiveHist | 34.4 | <0.001 |
| USC-INLA | 44.0 | <0.001 |
| kansas-bayesian | 51.2 | <0.001 |

Mean score differences and associated one-sided *p*-values for improvement of county-level climate-informed WNV forecast compared to each model analyzed within the 2022 CDC WNV Forecasting Challenge.

***Table S2: Improvements in Weighted Interval Score Components in the Univariate Climate-Informed Retrospective Forecast over the Historical Negative Binomial Model.***

| **NEON Region** | **Historical Negative Binomial Total WIS** | **Univariate Climate-InformedForecast Total WIS** | **Improvement in Total WIS** | **Improvement in Dispersion** | **Improvement in Overprediction** | **Improvement in Underprediction** |
| --- | --- | --- | --- | --- | --- | --- |
| Pacific Southwest | 0.34 | 0.26 | 0.08 | 0.00 | 0.05 | 0.04 |
| Desert Southwest | 0.44 | 0.39 | 0.05 | 0.02 | 0.05 | -0.02 |
| Northern Plains | 0.59 | 0.40 | 0.19 | 0.04 | 0.11 | 0.04 |
| Central Plains | 0.43 | 0.42 | 0.01 | -0.03 | 0.05 | -0.02 |
| Southern Plains | 0.7 | 0.47 | 0.23 | 0.08 | 0.11 | 0.03 |
| Prairie Peninsula | 0.44 | 0.31 | 0.13 | 0.02 | 0.08 | 0.03 |
| Ozarks Complex | 0.42 | 0.30 | 0.12 | 0.02 | 0.07 | 0.03 |
| Great Lakes | 0.51 | 0.46 | 0.05 | -0.01 | 0.08 | -0.03 |
| Southeast | 0.41 | 0.36 | 0.05 | 0.00 | 0.03 | 0.02 |
| Mid Atlantic | 0.38 | 0.31 | 0.07 | 0.01 | 0.03 | 0.03 |
| Northeast | 0.4 | 0.38 | 0.02 | -0.01 | 0.04 | 0.00 |
| Average | 0.46 | 0.37 | 0.09 | 0.01 | 0.06 | 0.01 |

Regional mean weighted interval scores from 2005-2022 for retrospective forecasts of the historical negative binomial model and the univariate climate-informed model, along with differences in total mean weighted interval score and the dispersion, overprediction, and underprediction components. Differences are computed as the historical negative binomial score minus the corresponding univariate climate-informed score so that improvements in weighted interval score components in the univariate model compared to the historical negative binomial baseline are positive.

***Table S3: Improvements in Weighted Interval Score Components in the Bivariate Climate-Informed Retrospective Forecast over the Historical Negative Binomial Model.***

| **NEON Region** | **Historical Negative Binomial Total WIS** | **Bivariate Climate-Informed Forecast Total WIS** | **Improvement in Total WIS** | **Improvement in Dispersion** | **Improvement in Overprediction** | **Improvement in Underprediction** |
| --- | --- | --- | --- | --- | --- | --- |
| Pacific Southwest | 0.34 | 0.27 | 0.07 | -0.01 | 0.04 | 0.04 |
| Desert Southwest | 0.44 | 0.41 | 0.03 | 0.03 | 0.05 | -0.04 |
| Northern Plains | 0.59 | 0.28 | 0.31 | 0.09 | 0.15 | 0.07 |
| Central Plains | 0.43 | 0.32 | 0.11 | 0.00 | 0.10 | 0.01 |
| Southern Plains | 0.7 | 0.48 | 0.22 | 0.07 | 0.11 | 0.03 |
| Prairie Peninsula | 0.44 | 0.29 | 0.15 | 0.02 | 0.09 | 0.04 |
| Ozarks Complex | 0.42 | 0.27 | 0.15 | 0.03 | 0.08 | 0.04 |
| Great Lakes | 0.51 | 0.46 | 0.05 | -0.01 | 0.09 | -0.03 |
| Southeast | 0.41 | 0.35 | 0.06 | -0.01 | 0.03 | 0.03 |
| Mid Atlantic | 0.38 | 0.25 | 0.13 | 0.02 | 0.05 | 0.05 |
| Northeast | 0.4 | 0.37 | 0.03 | -0.01 | 0.04 | 0.01 |
| Average | 0.46 | 0.34 | 0.12 | 0.02 | 0.07 | 0.02 |

Regional mean weighted interval scores from 2005-2022 for retrospective forecasts of the historical negative binomial model and the bivariate climate-informed model, along with differences in total mean weighted interval score and the dispersion, overprediction, and underprediction components. Differences are computed as the historical negative binomial score minus the corresponding univariate climate-informed score so that improvements in weighted interval score components in the bivariate model compared to the historical negative binomial baseline are positive.

***Table S4: Pairwise Comparison of Forecast Skill.***

| **NEON Region** | **Likelihood of Univariate Forecast Outperforming Historical Benchmark (%)** | **Likelihood of Bivariate Forecast Outperforming Historical Benchmark (%)** | **Likelihood of Bivariate Forecast Outperforming Univariate Forecast (%)** |
| --- | --- | --- | --- |
| Pacific Southwest | 96.81 | 93.74 | 0.29 |
| Desert Southwest | 85.04 | 70.51 | 24.95 |
| Northern Plains | 99.62 | 99.98 | 98.82 |
| Central Plains | 58.14 | 94.68 | 98.09 |
| Southern Plains | 97.19 | 96.72 | 31.55 |
| Prairie Peninsula | 96.92 | 98.56 | 75.95 |
| Ozarks Complex | 97.37 | 99.49 | 86.33 |
| Great Lakes | 73.17 | 72.64 | 54.65 |
| Southeast | 80.63 | 83.58 | 66.96 |
| Mid Atlantic | 92.01 | 98.37 | 95.82 |
| Northeast | 66.08 | 72.58 | 64.61 |
| National Aggregate | 99.09 | 99.81 | 98.7 |

Regional comparison of forecast skill between the retrospective forecasts for the univariate model, bivariate model, and historical negative binomial baseline. Values shown are the percentage of paired bootstrapped comparisons in which one model in the described pairing outperformed the other (model pairings described in column headers).

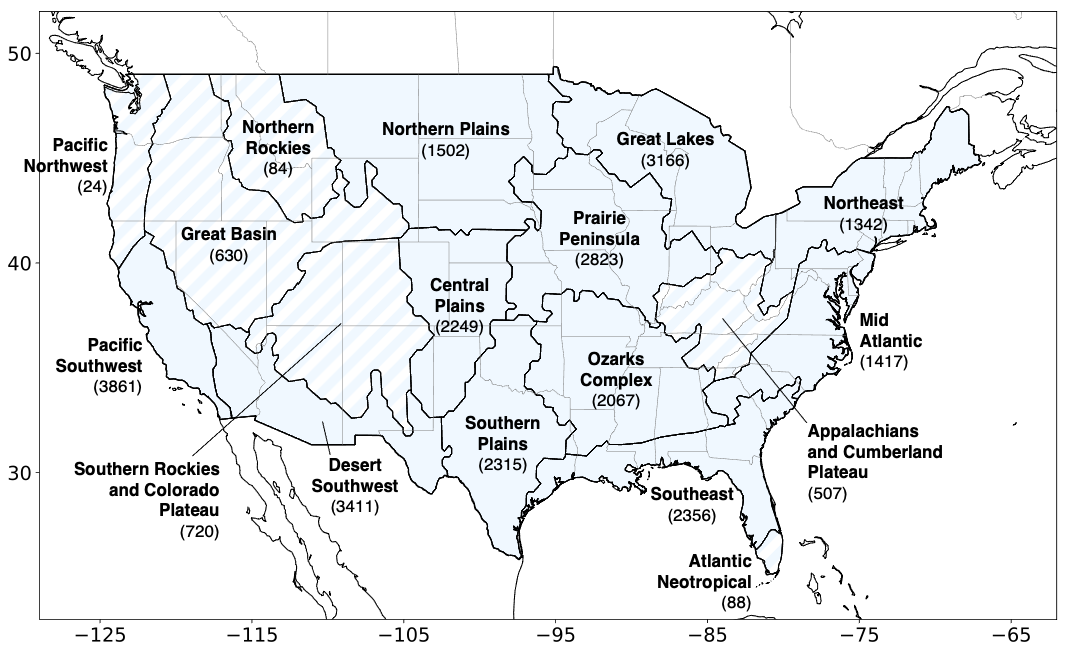
***Figure S1: NEON Regions with WNND Caseload from 1999-2022.*** A map of National Ecological Observatory Network (NEON) regions in the continental U.S. and the number of WNND cases reported to CDC ArboNET from 1999-2022. Regions with hatching were excluded from methodology here due to their relatively low number of WNND cases (< 1,000 WNND cases).

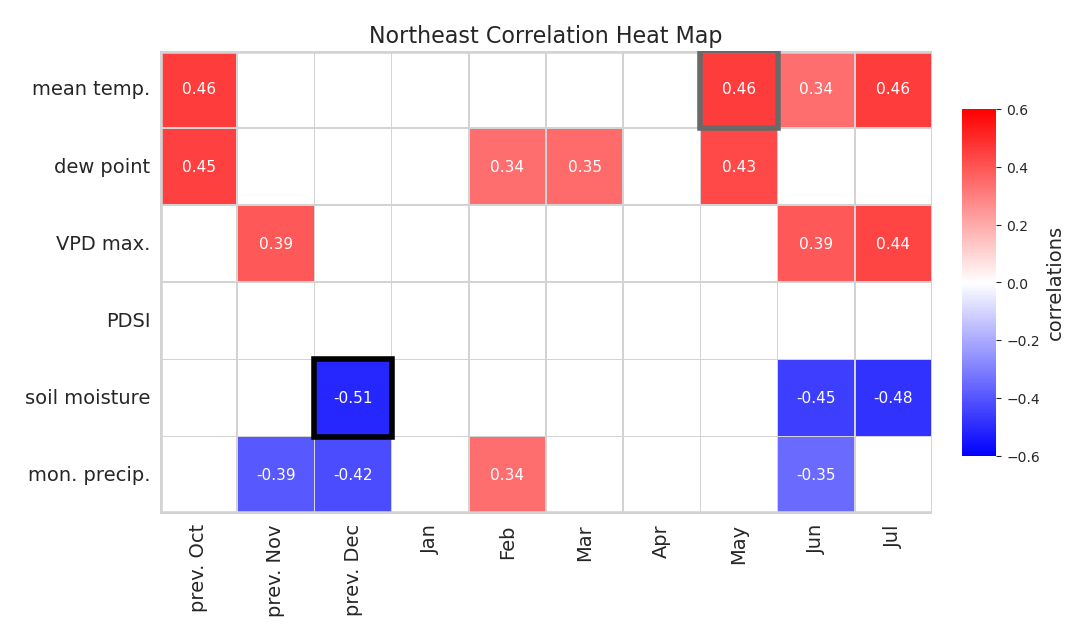
***Figure S2: Correlations between Monthly Climate Factors and Annual West Nile Virus Neuroinvasive Disease in the Northeast NEON Region.*** Each square shows the Pearson correlation (annotated red-blue fill) between annual WNND and the monthly climate factor represented by the given square for the Northeast NEON region. For example, the top right square represents the relationship between July mean temperature and annual WNND of the concurrent year (note that October–December columns represent correlations between climate conditions of the previous year with annual WNND as concurrent year climate conditions would have minimal impact on annual WNND totals given the strong August–September peak in WNV). Correlations between ±0.32 (*p*-value ~0.2) are masked to highlight stronger climate-WNND relationships. The regional primary and secondary climate variables selected for the bivariate WNV forecast model are outlined in thick black and gray, respectively.

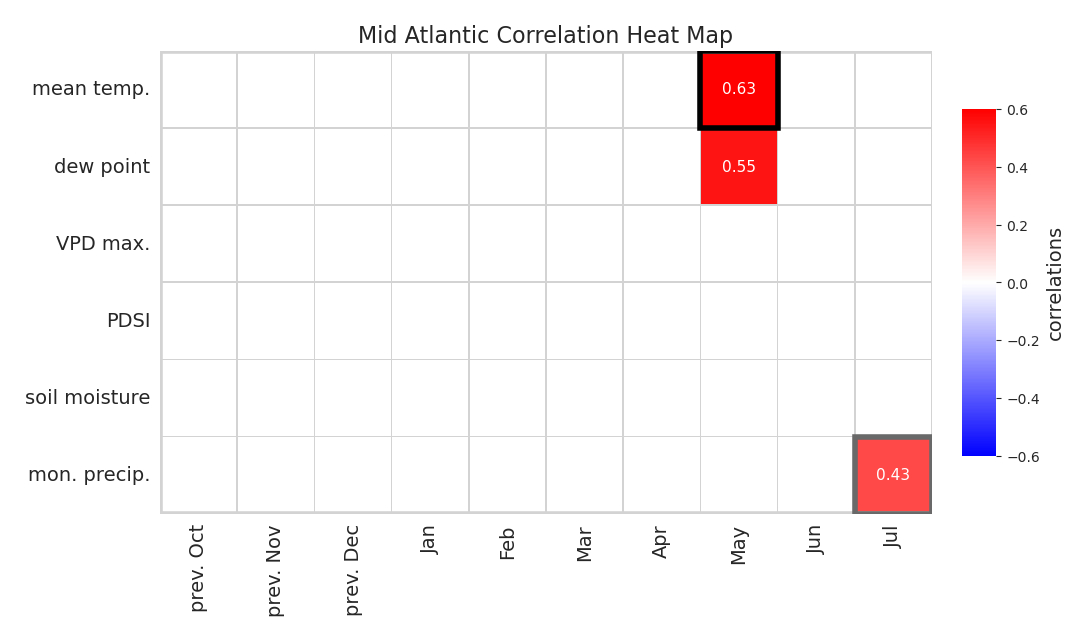
***Figure S3: Correlations between Monthly Climate Factors and Annual West Nile Virus Neuroinvasive Disease in the Mid Atlantic NEON Region.*** Same as Figure S2 but for the Mid Atlantic NEON region.

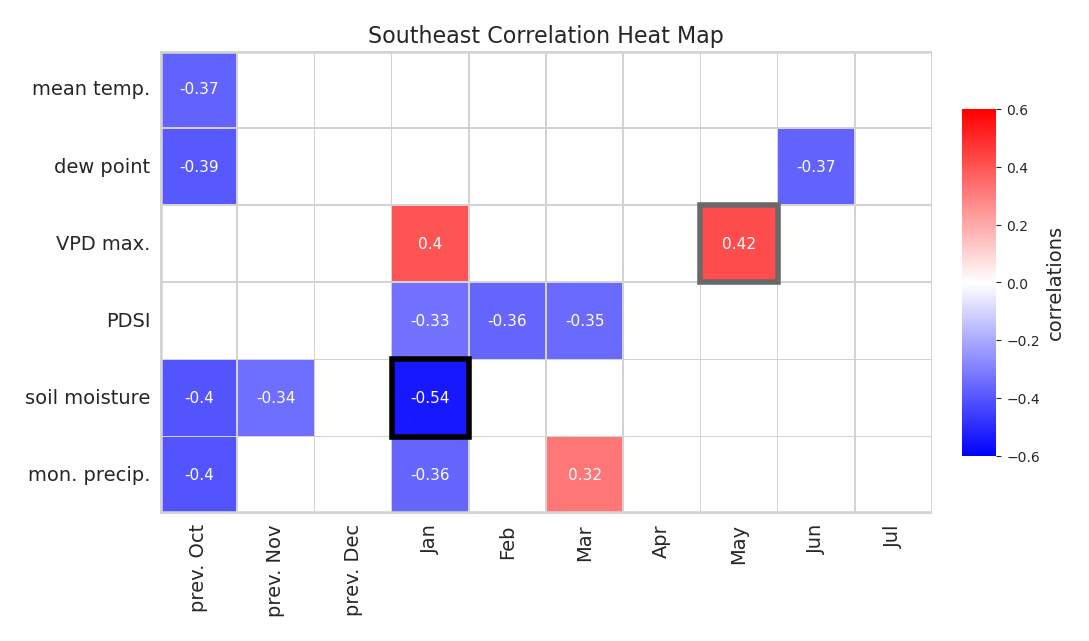
***Figure S4: Correlations between Monthly Climate Factors and Annual West Nile Virus Neuroinvasive Disease in the Southeast NEON Region.*** Same as Figure S2 but for the Southeast NEON region.

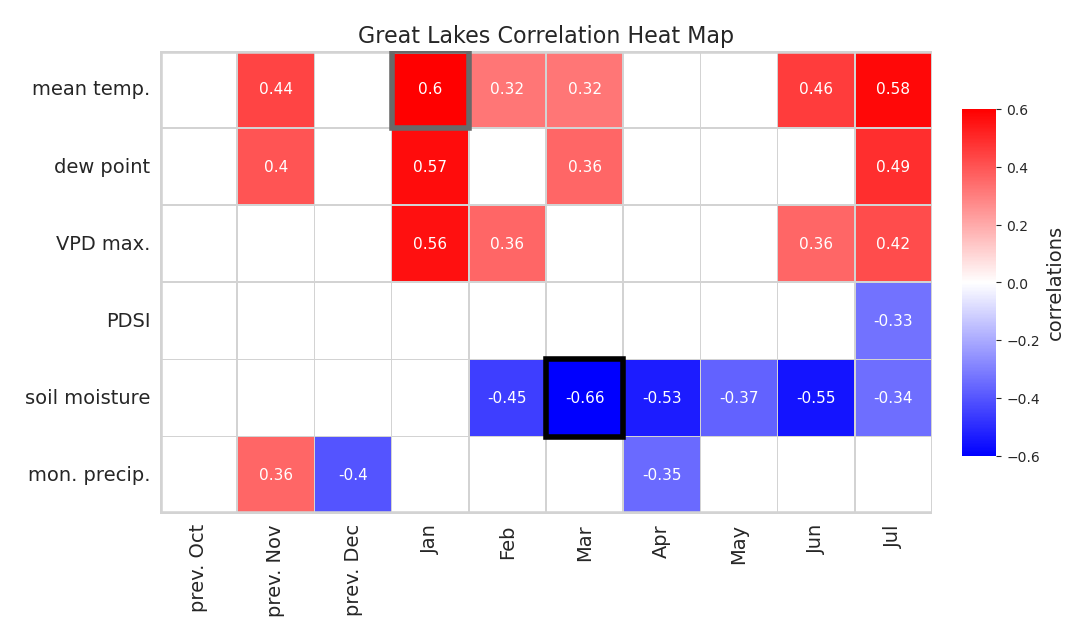
***Figure S5: Correlations between Monthly Climate Factors and Annual West Nile Virus Neuroinvasive Disease in the Great Lakes NEON Region.*** Same as Figure S2 but for the Great Lakes NEON region.

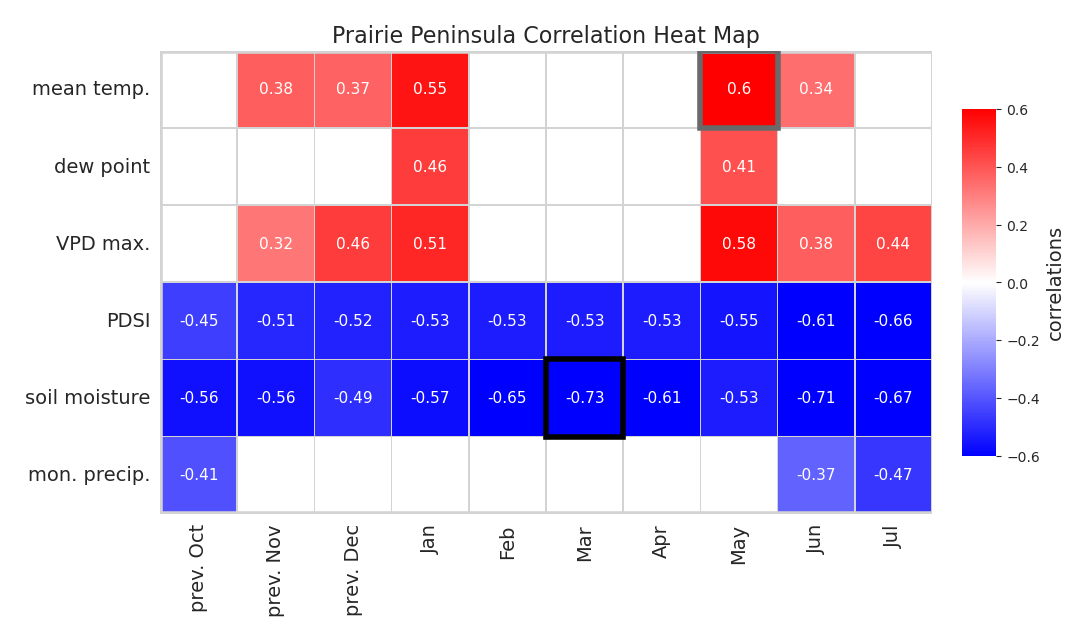
***Figure S6: Correlations between Monthly Climate Factors and Annual West Nile Virus Neuroinvasive Disease in the Prairie Peninsula NEON Region.*** Same as Figure S2 but for the Prairie Peninsula NEON region.

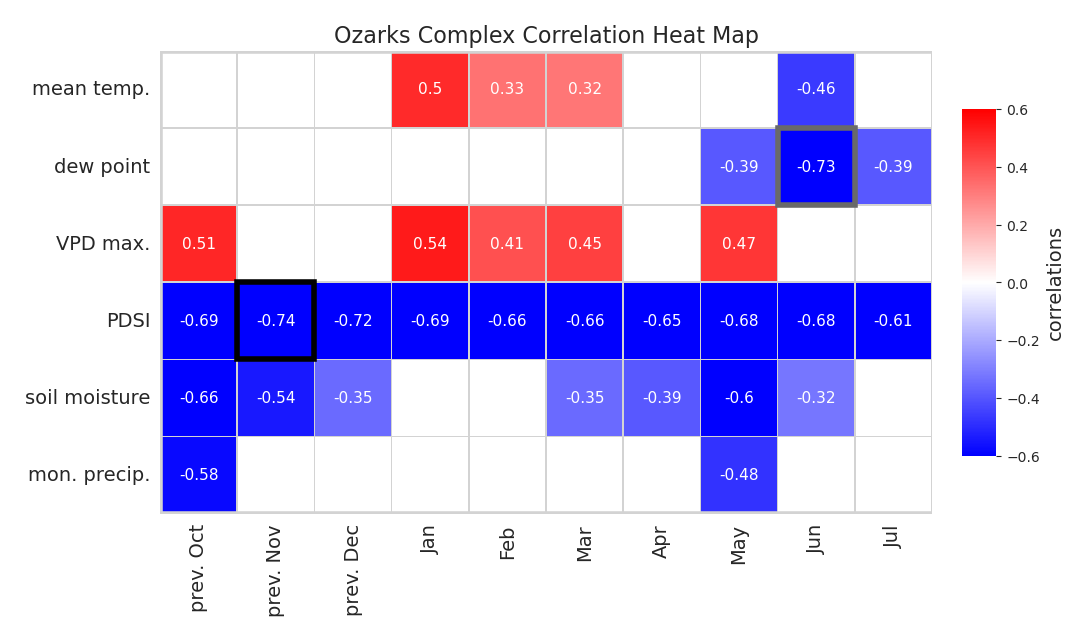
***Figure S7: Correlations between Monthly Climate Factors and Annual West Nile Virus Neuroinvasive Disease in the Ozarks Complex NEON Region.*** Same as Figure S2 but for the Ozarks Complex NEON region.

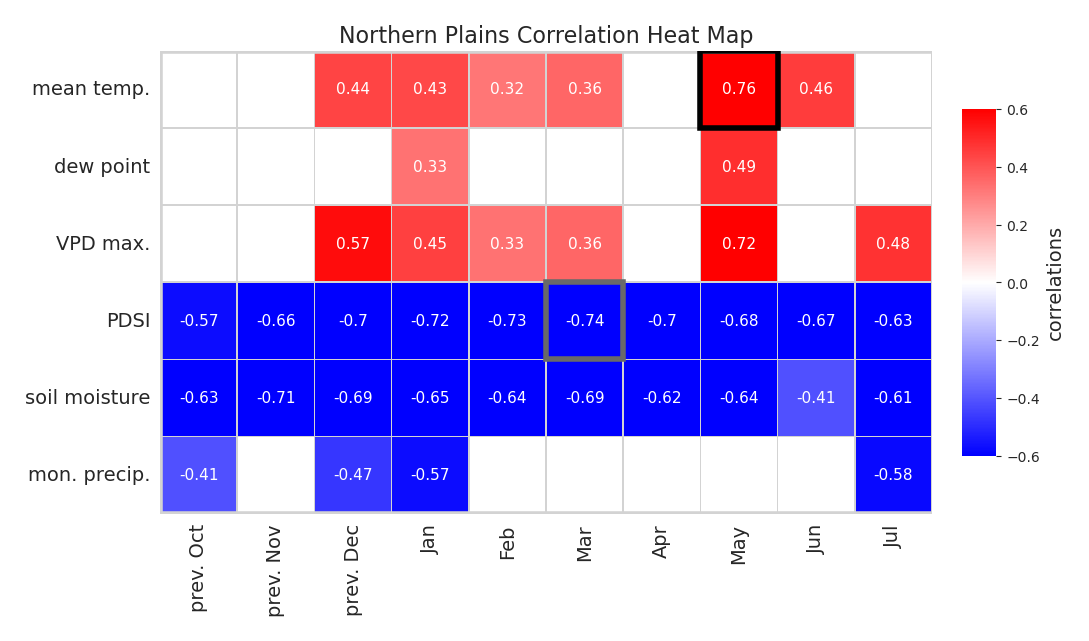
***Figure S8: Correlations between Monthly Climate Factors and Annual West Nile Virus Neuroinvasive Disease in the Northern Plains NEON Region.*** Same as Figure S2 but for the Northern Plains NEON region.

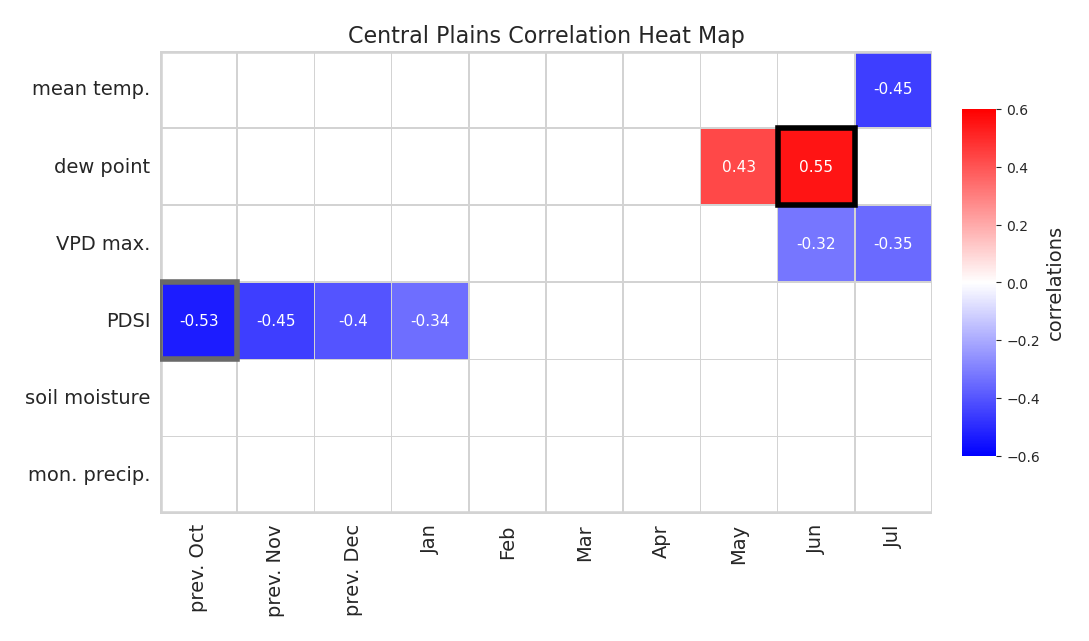
***Figure S9: Correlations between Monthly Climate Factors and Annual West Nile Virus Neuroinvasive Disease in the Central Plains NEON Region.*** Same as Figure S2 but for the Central Plains NEON region.

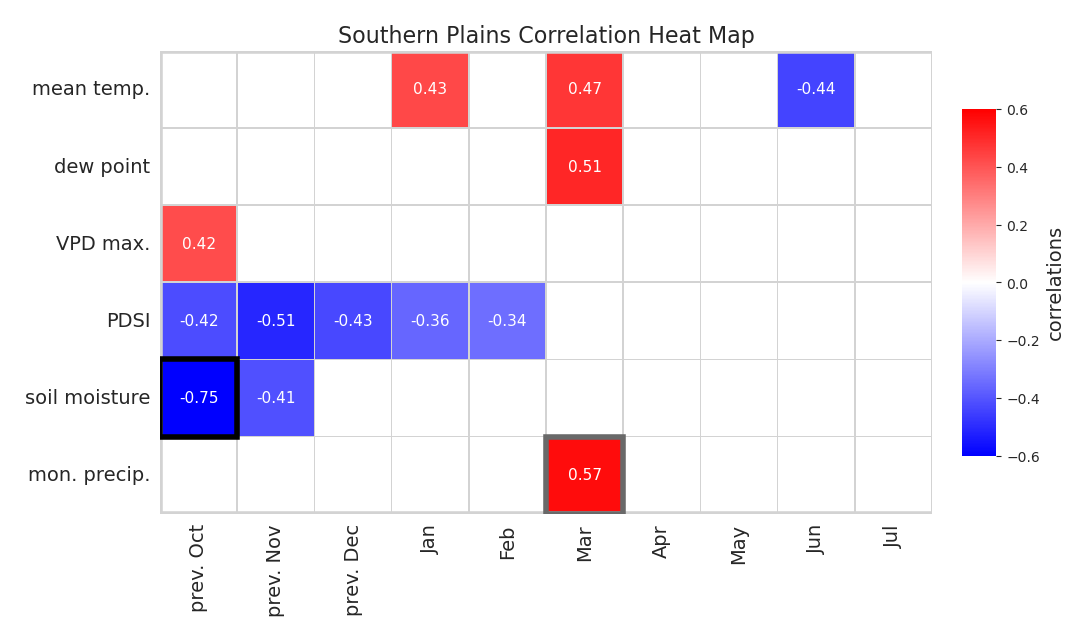
***Figure S10: Correlations between Monthly Climate Factors and Annual West Nile Virus Neuroinvasive Disease in the Southern Plains NEON Region.*** Same as Figure S2 but for the Southern Plains NEON region.

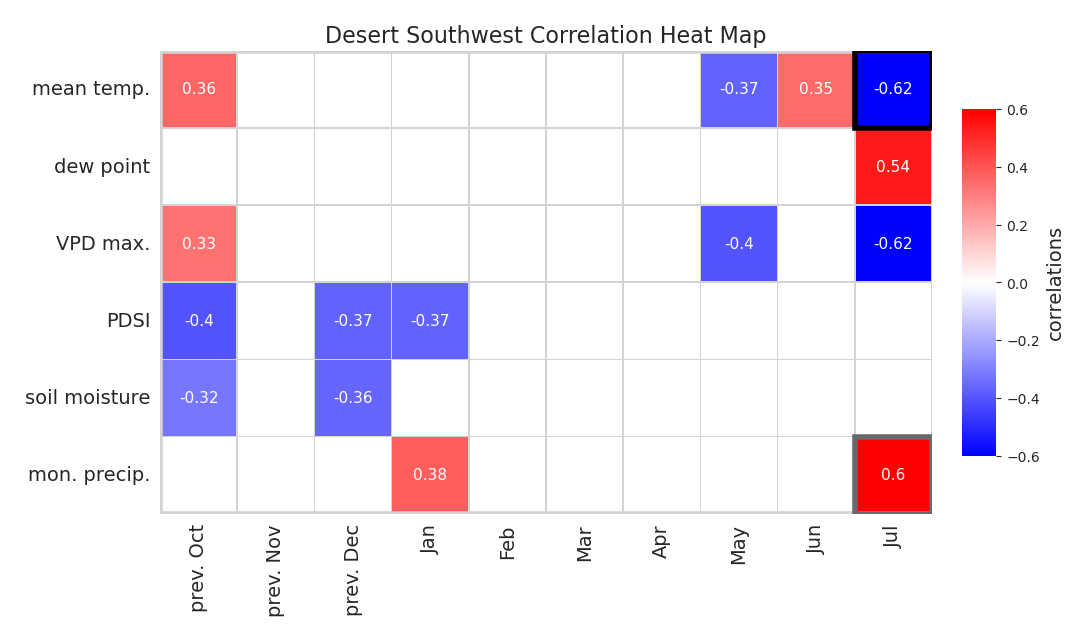
***Figure S11: Correlations between Monthly Climate Factors and Annual West Nile Virus Neuroinvasive Disease in the Desert Southwest NEON Region.*** Same as Figure S2 but for the Desert Southwest NEON region.

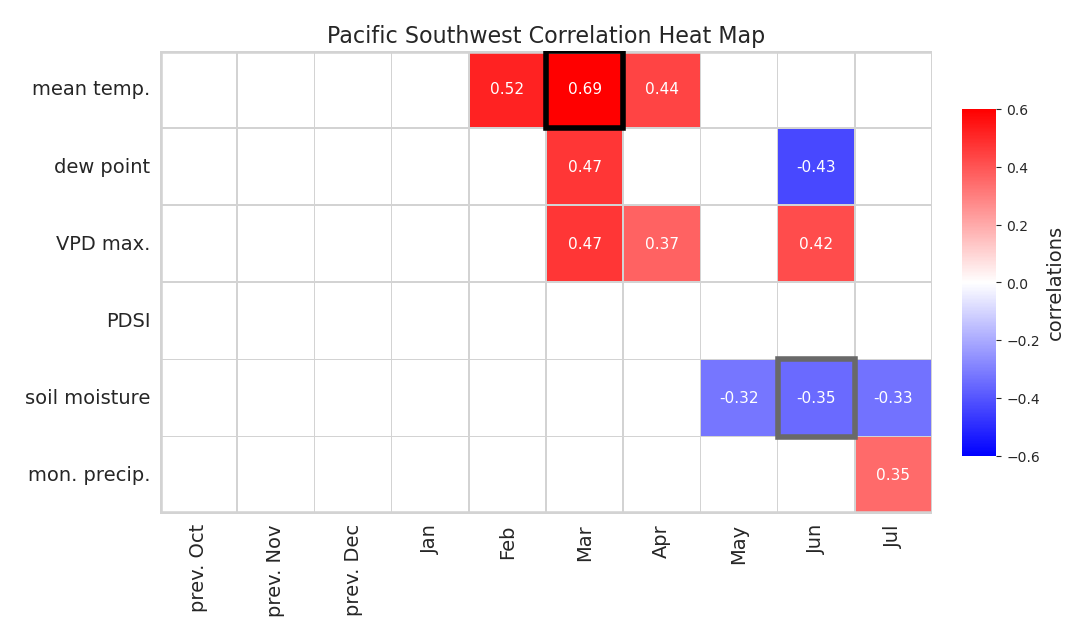

***Figure S12: Correlations between Monthly Climate Factors and Annual West Nile Virus Neuroinvasive Disease in the Pacific Southwest NEON Region.*** Same as Figure S2 but for the Pacific Southwest NEON region.

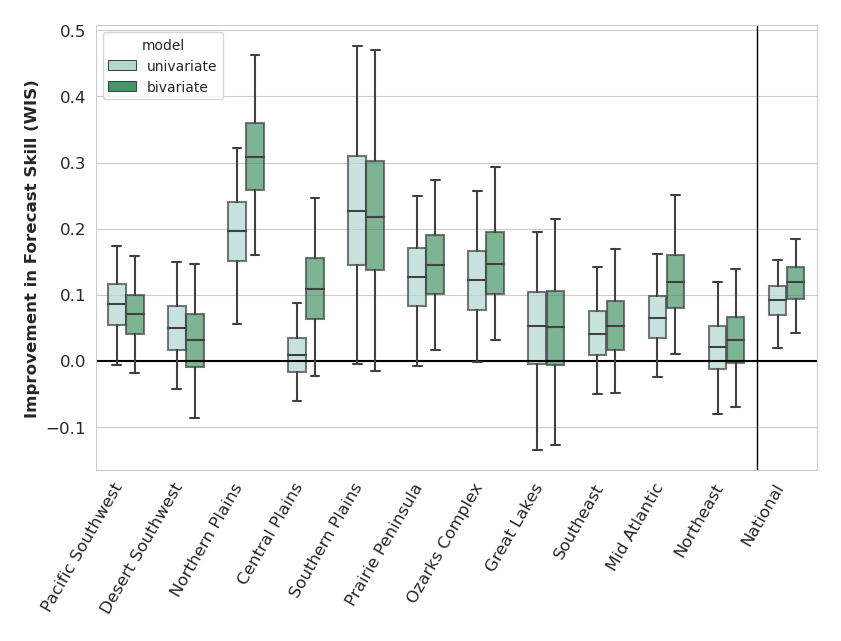

***Figure S13: Weighted Interval Score Improvement in Regional Forecast Skill of Climate-Informed Forecast Models Compared to Historical Negative Binomial Benchmark.*** Box plots of regional and nationally aggregated differences in retrospective forecast skill between univariate (light green) and bivariate (dark green) climate-informed forecast models and historical negative binomial distribution for 2005–2022. Score improvements are calculated as the climate-informed forecast model weighted interval scores less the historical negative binomial weighted interval scores. These differences are then inverted so that above zero indicate regions where forecast skill is better in the climate-informed forecast model. Box encompasses interquartile range and whiskers depict 95% confidence intervals as determined by bootstrapping. NEON regions are listed from West to East
